# Embodied Interoception Questionnaire (Intero-10): development, validation, and application in people with chronic pain

**DOI:** 10.1101/2025.09.12.25335640

**Authors:** Ana Mércia Fernandes, Vinicius Deniz de Oliveira, Samantha K. Millard, Carolina Campos dos Santos, Jorge Alberto Martins Pentiado Júnior, Marcell Maduro Barbosa, Pedro Nascimento Martins, Suzana Curcino Nogueira, Maercio Maia Alves, Pedro Henrique Martins da Cunha, Danielle Cristina Fonseca, Luccas Soares LaFerreira, Paulo Roberto Santos-Silva, Nayara Salgado Carvalho, Vinícius Carlos Iamonti, Celso R. F. Carvalho, Camila Squarzoni Dale, Gabriel Taricani Kubota, Lin Tchia Yeng, Manoel Jacobsen Teixeira, Abrahão Fontes Baptista, Interoception Study Group, Daniel Ciampi de Andrade

## Abstract

Embodied interoception refers to the perception of the body’s state and is a multidimensional cognitive process. It is proposed that the experience of pain also feeds interoceptive networks with information from the state of the body, and the subjective experience of pain would be influenced by an individual’s trait embodied interoceptive profile. Here we developed and validated the Intero-10, a questionnaire designed to specifically evaluate embodied interoception based on trait interoceptive channels. Healthy adults (n=381) and people with neuropathic pain (n=86) were enrolled. The relationship between trait and state interoceptive responses was examined during experimentally evoked interoceptive psychophysics tasks, which were specific for each interoceptive channel. During these tests, embodied interoceptive state scores only correlated with trait ones for unpleasantness (*P*<0.05), but not for intensity rating, suggesting that the predicted negative valence of lived experience provided lower prediction errors than intensity estimations. The Intero-10 final version included embodied interoceptive perception intensity (heartbeat, heat, itching, dyspnea, sleep, muscle fatigue, and anguish) and unpleasantness (heartbeat, dyspnea, and nausea) categories. Intero-10 demonstrated adequate content validity, good internal consistency (Cronbach’s alpha=0.81), good reliability (>0.75), and a single-factor structure. Patients with neuropathic pain filled in the Intero-10 alongside traditional pain, mood, sleep, and quality of life assessments. Embodied interoception scores correlated with mood, and quality of life, and partially mediated the correlation between pain interference and quality of life (β=-0.0093). The specific assessment of embodied interoceptive channels may broaden our current assessment of people with chronic pain and of those at risk to develop it.

## Introduction

In 1906 Charles Sherrington framed the concept of interoception as being “sensations originating from within the body, particularly from the viscera”.(Ceunen et al., 2016; Sherrington, 1906) It is currently defined as the perception of the body’s state.(Ceunen et al., 2016; A. D. Craig, 2002) Interoceptive inputs play a central role in processes such as allostasis - the body’s capacity to predict and optimize the body’s energy use.(Barrett & Simmons, 2015; Katsumi et al., 2022) Interoceptive signal processing shares many pathways with the nociceptive system. Both involve spino-thalamo-cortical tracts conveying discriminatory sensory inputs from small-diameter afferent fibers located in both somatic and visceral tissues through the thalamic ventromedial posterior nucleus up to the granular posterior aspect of the insula.(Ceunen et al., 2016; A. D. Craig, 2002; Fermin et al., 2021) In the model of interoceptive predictive coding, external or internal sensory inputs update internal allostatic predictions originating from visceromotor cortical areas, allowing for the measurement of the extent to which internal predictions (priors) match incoming sensory inputs. As discrepancies between priors and sensory inputs occur, prediction error is computed.(Barrett & Simmons, 2015; Seth et al., 2011) Within this coding framework, prediction errors are minimized either by changing the gain of sensory inputs, by updating predictions about the world/body, or by (en)acting upon the body/environment in order to generate novel sensory inputs that better align with internal predictive models.(Seth, 2013; Seth et al., 2011) It has been proposed that the experience of pain would also feed interoceptive networks with information from the state of the body,(Kiverstein et al., 2022) and the subjective experience of pain would be influenced by an individual’s trait embodied interoceptive profile,(Di Lernia et al., 2016) and by momentary processes influencing their interoceptive experiences.(Pollatos et al., 2012)

Disruptions in interoceptive processes have been found in chronic pain(Di Lernia et al., 2016, 2020; Horsburgh et al., 2024; Sedley et al., 2024) and psychiatric disorders.(Critchley & Garfinkel, 2017; K. Friston, 2010; K. J. Friston, 2017; Nord & Garfinkel, 2022; Seth et al., 2011) Despite these reported interactions, the interplay between pain and embodied interoceptive experiences has rarely been explored, mainly due the lack of specific tools to specifically detangle embodied interoceptive perceptions from other interoceptive constructs such as attention, accuracy, awareness, and emotional awareness.(Gabriele et al., 2022; Mehling et al., 2012; Murphy et al., 2020; Schandry, 1981) While several tools address these later domains, most were not designed to specifically assess the embodied experience—the direct perception of internal bodily sensations, and none were specifically designed to be used in people with chronic pain.

In the present study, the Intero-10 questionnaire was developed to assess trait embodied interoceptive channels (self-perceived usual experience of embodied interoception). Its items were correlated with state (momentary lived experience) perceptions evoked by experimentally probing each of interoceptive channels. Following item-reduction, a full validation process allowed for the report of normative values and its use in people with chronic pain.

## Methods

### Study design

This was a study to develop and validate an embodied interoception questionnaire including five steps (Fig. 1). Step 1 - a systematic literature review was conducted to list the interoceptive channels previously explored in psychophysics setups; Step 2 - an embodied interoceptive trait proto-questionnaire was developed with channels selected in step 1. The “trait” proto-questionnaire underwent face and content validities, and then it was answered by a group of healthy participants; Step 3 - healthy volunteers who had previously filled in the embodied interoceptive trait proto-questionnaire were invited to participate in one or more of 16 interoceptive psychophysics setups (interoceptive “state”). For example, the sensation of thirst was evoked by the consumption of a standardized dose of salted potato chips (Brannigan et al., 2015), then the thirst perception intensity and thirst unpleasantness levels were measured; Step 4 - a correlation analysis was conducted between responses to the embodied interoceptive trait proto-questionnaire (step 2), and both interoceptive perception intensity and interoceptive unpleasantness reported during the interoceptive state test of each channel (step 3); Step 5 - the questionnaire was formally validated using psychometric analyses; a latent variable modeling approach was used for the detection and removal of redundant items of the proto-questionnaire, and creating the Embodied Interoception Questionnaires (Intero-10); additionally, patients with chronic neuropathic pain answered the Intero-10 and other traditional pain, mood and quality of life assessment scales and questionnaires. This was used for the exploration of how the Intero-10 relates to other chronic pain constructs, as well as allowing for the delineation of discriminant validity and mediation analyses.

**Figure 1.**
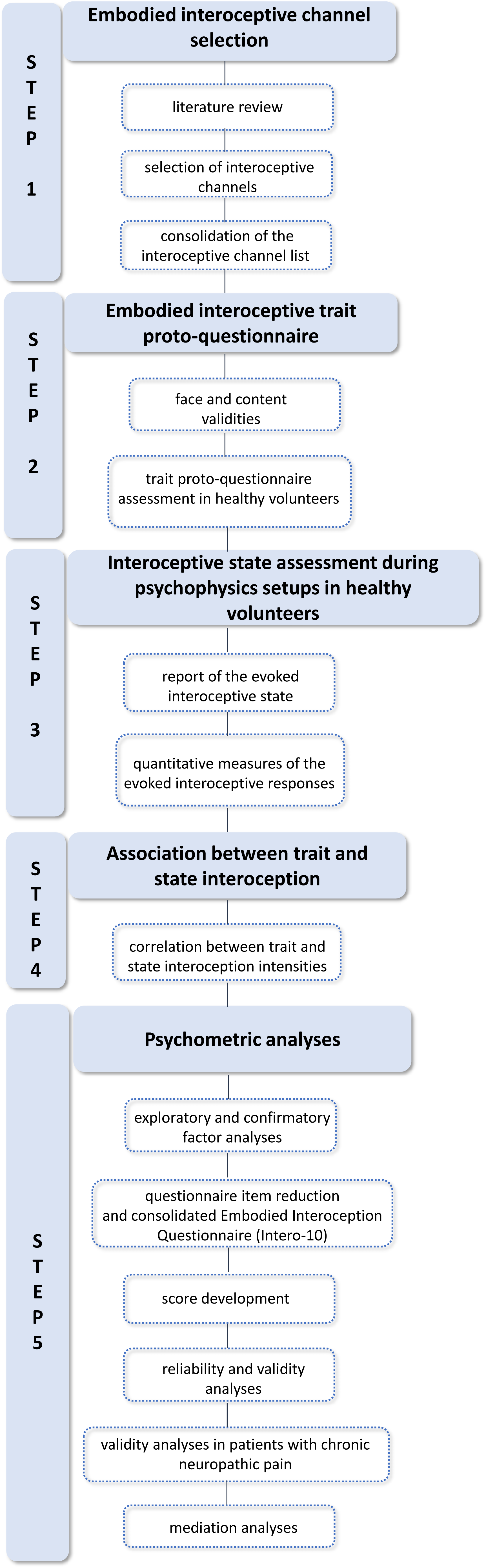
Overall study design flowchart. Step 1 involved selecting embodied interoceptive channels. Step 2 focused on developing the embodied interoceptive trait proto-questionnaire, assessing face and content validity, and applying it to a healthy participant group. Step 3 evaluated state interoception through psychophysics setups in healthy adults. Step 4 analyzed the association between trait and state interoception. Step 5 involved psychometric evaluation, including reducing questionnaire items.

### Ethics

The Institutional Local Ethics Review Board (#69685817.7.0000.0068) of the University of São Paulo, Brazil, approved this study, with all volunteers providing informed consent before the study protocol was implemented. The sampling was non-probabilistic convenience sampling.

### Setting and data collection

This study was conducted at the Department of Neurology at HC-FMUSP (Clinics Hospital of the School of Medicine at the University of São Paulo), and the recruitment of study volunteers started in 2018. All collected survey data were compiled into REDCap software^®^.(Harris et al., 2009, 2019)

### Participants

All volunteers (healthy and patients) answered the following socio-demographic data: (a) age in years; (b) gender (male, female); (c) ethnicity; (d) religion; (e) weight in kilograms; (f) height in meters; (g) body mass index (BMI) calculated as weight (in kilograms) over height squared (in meters); (h) comorbidity; (i) occupation; (j) marital status; and (k) smoking and drinking status.

### Healthy volunteers

Healthy people, who could read and write in Brazilian Portuguese language, aged between 18 and 59 years old, participated in steps 2 and 3. The sample size was based on having 10 subjects per variable or item, with a minimum of 100 subjects in the total sample.(Crocker & Algina, 1986; Tsang et al., 2017) Exclusion criteria were severe neuropsychiatric disorders such as severe depression, oncological diseases, acute or chronic infections, pregnancy, diabetic patients, patients with chronic pain, people with chronic alcoholism or substance abuse. Volunteers with any hearing, visual, physical, or cognitive impairment that would prevent their ability to respond to study questions were also excluded. At least 20 volunteers (equally distributed among sexes) were included in the evaluation of 16 different psychophysical tests (step 3). For each test, distinct interoceptive body experiences were established and applied specific inclusion criteria, which will be elucidated later in this study. Some volunteers participated in multiple psychophysical sessions.

For convergent validity in step 5, they answered the embodied interoceptive trait questionnaire and other interoceptive questionnaires: Multidimensional Assessment of Interoceptive Awareness (MAIA),(Mehling et al., 2012) Interoceptive Accuracy Scale (IAS),(Murphy et al., 2020) Interoceptive Confusion Questionnaire (ICQ),(Brewer et al., 2016; Goerlich, 2018) Body Perception Questionnaire (BPQ)(Cerritelli et al., 2021) and Toronto alexithymia scale (TAS-20).(Fortes et al., 2017)

### Patients with chronic neuropathic pain

In step 5, people with neuropathic pain were included based on the current International Association for the Study of Pain (IASP) definitions, and grading system as having definite neuropathic pain.(Raja et al., 2020) They were recruited from pain clinics of the Hospital das Clínicas, University of São Paulo. They answered the embodied interoception questionnaire and pain intensity using VAS, brief pain inventory (BPI),(Ferreira et al., 2011; Melzack, 1987) neuropathic pain symptom inventory (NPSI),(de Andrade et al., 2011) sleep scale from the Medical Outcome Study (MOS),(Hays et al., 2005) hospital anxiety and depression scale (HADS),(Zigmond & Snaith, 1983) and short-form 6 dimensions (SF-6D).(Campolina et al., 2011)

### Description of steps

#### Step 1 (Embodied interoceptive channel selection)

In this step, the aim was to list embodied interoceptive channels that were explored during an experimental session (psychophysics setups). For that, a literature review was conducted aimed at identifying interoceptive channels and their assessment models. The following search terms was executed in the PubMed database: “((interoception OR interoceptive OR body awareness OR interoceptive awareness OR visceral sensations) AND (visceral pain OR thirst OR heartbeat perception OR hunger OR heat OR cold OR itching OR sleep OR fatigue OR nausea)) AND (questionnaire OR scale OR rating OR instrument OR inventory)”. All fields of search on PubMed were included, spanning the title, abstract, and the main body of the text. Additionally, other relevant papers related to interoceptive sensations were incorporated, which were identified by authors of this study. Selection criteria for interoceptive channels were: experimental test models previously utilized for evaluating interoceptive channels, feasible execution, which did not cause suffering to the volunteer, and published protocols. Feasibility was defined as the practicality of measuring the interoceptive channel in a controlled setting, considering factors such as the availability of reliable equipment, the participant’s ability to tolerate the procedures, and the time required for data collection. Additionally, the selected channels had to demonstrate consistent and replicable responses in prior studies, ensuring that they could provide meaningful responses into interoceptive stimuli. Subsequently, the list of embodied interoceptive channels was used in step 2 in healthy volunteers and step 3 in which chosen protocols were reproduced or adapted in this study.

#### Step 2 (Embodied interoceptive trait proto-questionnaire)

A preliminary questionnaire was developed: the embodied interoceptive trait proto-questionnaire. The term “trait” is used here as a self-perceived usual experience, based on previous descriptions of interoception studies.(Garfinkel et al., 2015) The embodied interoceptive trait proto-questionnaire included an introductory part with instruction for fulfillment and two questions for each interoceptive channel. These questions were presented on a horizontal visual analogue scale (VAS), ranged from 0 to 100 mm, in which terms “I do not perceive it at all” and “I perceive it at the maximum possible intensity” were used to assess the interoceptive perception intensities in daily life and “I am not bothered at all” and “It bothers at the maximum possible intensity” to assess the level of unpleasantness.

#### Face and content validity

Face and content validity are essential concepts in psychometrics, used to evaluate the quality of a test or measurement instrument. Face validity refers to the extent to which a test or measurement instrument appears to measure what it claims to measure, based purely on superficial examination. It does not involve statistical tests but rather an intuitive and subjective judgment about the instrument’s relevance made by non-experts, participants, or stakeholders who evaluate whether the items on a test seem relevant and appropriate for the construct being assessed. High face validity is useful for ensuring test-takers’ confidence and motivation, as they are more likely to engage meaningfully with a test if it seems to be measuring what it claims to.(Nevo, 1985) Content validity ensures that a questionnaire includes all the entire domain of the construct it aims to assess, while excluding irrelevant items(Haynes et al., 1995) A panel of five experts (LL, IR, MJT, SCN, and FG) were invited to evaluate the proto-questionnaire, based on their clinical expertise and professional backgrounds in clinical assessment, and/or their experience in questionnaire’s development.(Yaddanapudi & Yaddanapudi, 2019) They were informed the questionnairés items were chosen based on previously explored psychophysics setups reported in the scientific literature. This multidisciplinary group, consisting of neuroscientists, neurosurgeons, physiotherapists, and neurologists, participated in a Delphi method process,(Vernon, 2009) which involved up to five rounds of email surveys/communications. They reviewed both the introduction and individual items of the questionnaire, assessing them qualitatively and quantitatively. A semantic analysis was conducted to determine the clarity, intelligibility, and relevance of the questions. During the qualitative analysis, experts provided their feedback on items and the terminology used in the questionnaire, and whether the embodied interoceptive trait proto-questionnaire items were pertinent and essential to the scale. They also had the opportunity to suggest modifications, exclusions, or additions. For the quantitative analysis, the content validity coefficient (CVC)(Hernandez-Nieto, 2002) was calculated to measure the degree of agreement between experts. Using a Likert-type scale (1 = unclear/irrelevant, 2 = needs major revision, 3 = clear/relevant with minor changes, 4 = absolutely clear/relevant), the CVC was calculated for each item. The total CVC (CVC*t*) of each item was computed by subtracting the standard error (Pe*i*), which is obtained by the ratio of 1 over the number of judges to the number of judges, from the initial CVC (CVC*i*): CVC*t =* CVC*i* - Pe*i,* according to the following formula:

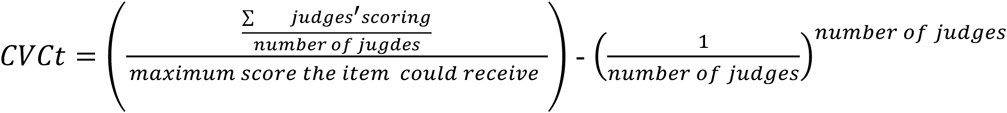

Scores range from 0 to 1, with values of 0.80 or higher indicating a high-quality item.(Hernandez-Nieto, 2002) Items scoring closer to 1 were either retained or modified based on the consensus of the experts.

Following face and content validity, a pilot test was conducted with a representative sample population (n=20) to identify any remaining issues related to item clarity, ambiguity, or relevance that were not apparent in the expert review process.(Hertzog, 2008; van Teijlingen & Hundley, 2002) After the pilot test, the embodied interoceptive trait proto-questionnaire was completed by healthy volunteers.

Additionally, the Embodied interoceptive trait proto-questionnaire was translated from Portuguese to English following established Guidelines.(Guillemin et al., 1993) This process involved: 1) Translation: three independent, and fluent in the idioms translators participated in this phase, identifying errors and divergent interpretations of ambiguous items in the original; 2) Back-translation: this phase produced as many back-translations as translations to help improve the quality of the English version, and 3) Committee review: a multidisciplinary group of neuroscientists, psychologists, and neurologists compared source and final versions, resolving discrepancies, modifying instructions, modifying/rejecting inappropriate items, and ensuring cross-cultural equivalence and comprehensibility.

#### Step 3 (Interoceptive state assessment during psychophysics setups)

The objective of this step was to assess the interoceptive state rating by healthy individuals during the actual experience of each of the interoceptive channels during 16 different psychophysics settings. In this context, the term “state” (momentary lived experience) refers to the interoceptive perception experienced by healthy volunteers when exposed to stimuli capable of eliciting interoceptive sensations. Invitations were extended to a minimum of 20 healthy volunteers who had previously completed the embodied interoceptive trait proto-questionnaire to participate in one or more of interoceptive psychophysics setup.

In total, 16 out of the 17 interoceptive channels included in the embodied interoceptive trait proto-questionnaire were assessed in this step. Similarly, they answered questions about the intensity of embodied interoceptive perception evoked by the experience as well as their unpleasantness level associated with these perceptions using a visual analogue scale (VAS). Additionally, biological and/or quantitative variables related to each interoceptive channel were recorded whenever it was possible. For each interoceptive channel, a specific psychophysics setup was adopted, as described below. Detailed information about each psychophysical setup is available in Supplementary Text S1.

i. Deep pressure pain: deep pressure hyperalgesia was measured over the thenar eminence of the right hand based on a 1.3x pressure pain threshold for 5 seconds and measured using VAS.(Petrini et al., 2015)
ii. Visceral pain perception: visceral pain intensity caused by uterine cervix clamp was obtained during routine DIU insertion using VAS.(Lanzola & Ketvertis, 2023) The interval of visceral pain in seconds (caused by pressure on the cervix during IUD implantation) was measured in this assessment.
iii. Heartbeat perception: The Heartbeat Detection Tracking Task was used to measure the heartbeat perception.(Schandry, 1981) After this protocol, volunteers were asked how well they believed they had counted their heartbeats and how unpleasant it was to perform this task. This evaluation is related to interoceptive awareness, an aspect of interoception assessed via metacognitive judgments of interoceptive accuracy.(Garfinkel & Critchley, 2013)
iv. Hunger perception: participants rated their hunger perception intensity and unpleasantness level using VAS after fasting, and after consuming a test meal. The biological measure of the interoceptive sensation of hunger was the assessment of ghrelin and leptin hormone levels in the blood plasma.(Malin et al., 2020)
v. Thirst perception: volunteers consumed salty potato chips to induce thirst according to an adapted protocol.(Brannigan et al., 2015) They answered the thirst perception intensity and unpleasantness level using VAS at 3 different moments. The amount of water consumed was the quantitative measure of this assessment.
vi. Environmental temperature perception: volunteers were exposed to rooms that were hot (an average of thirty-eight degrees Celsius) or cold (an average of sixteen degrees Celsius), then the ear temperature of the participants was measured. This protocol was adapted from a previous study.(Trezza et al., 2015)
vii. Skin itching perception: skin prick testing (SPT) with histamine dihydrochloride (10 mg/ml in 50% glycerin solution with 0.4% phenol) was used to induce itching according to adapted protocol(Heinzerling et al., 2013) in an area of 25 cm^2^ in the left forearm. After 1, 5, and 10 minutes of the test, the itching perception intensity and unpleasantness that the test caused were calculated using VAS.
viii. Dyspnea perception: inspiratory endurance test was used to assess the dyspnea perception using a PowerBreathe (Medical KH2^®^, IMT Technologies Ltd, Birmingham, UK).(Cahalin & Arena, 2015) Volunteers assessed their dyspnea perception intensity and unpleasantness using VAS.
ix. Sleep duration perception: volunteers used a sleep self-reported questionnaire and an actigraph (Act Trust^®^, Condor Instruments, São Paulo, Brasil) to register sleep parameters commonly estimated in sleep logs. The aim of this assessment was to measure and correlate the self-reported total sleep time (TST), and time in bed (TIB) with data obtained from the actigraph.
x. Muscle fatigue perception: volunteers underwent exercise testing on an ergometric bicycle (bike, Cybex^®^, USA) using the modified Astrand protocol to assess muscle fatigue.(Åstrand et al., 2003) For quantitative variable analysis, VO2máx (ml/kg/min) was used at ventilatory thresholds 2 (VT2). Furthermore, muscle fatigue perception intensity and unpleasantness using VAS was registered.
xi. Bladder fullness perception: after spontaneous urination, participants were asked to drink 500 mL of water every half hour and to report when they reached three standard cystometric urgency thresholds: first sensation, first desire to urinate and maximum capacity.(Abrams et al., 2002) Bladder volume was measured with three-dimensional ultrasonographic measurements, and participants assessed bladder fullness perception intensity and unpleasantness using VAS.
xii. Respiratory frequency and gastric fullness perception: participants underwent the Electrogastrography (EGG) examination and a Walter Load Test (WLT) protocol(Chen et al., 2004; Jones et al., 2003) for induction of gastric fullness. They assessed gastric fullness perception intensity and unpleasantness using VAS. The respiratory strap placed in the lower thoracic region registered the respiratory frequency during the next 30-minutes while the participant performed the mental tracking task. This task was the same as the protocol used in evaluating heartbeat perception,(Pollatos et al., 2012; Schandry, 1981) however participants were asked to mentally count their respiratory frequency. Afterwards, just as heartbeat evaluation, the evaluator asked how well they believed they had counted their respiratory frequency and how unpleasant it was to perform this task. This evaluation is also related to metacognitive awareness of interoceptive accuracy.
xiii. Nausea and anguish perception: specialist committee experienced in validating a set of emotional facial expressions selected a 1-minute-and-40-second video from a group of videos that could evoke the sensation of nausea. Additionally, the committee selected a 2-minute-and-30-second video that could induce the sensation of anguish. Volunteers watched nausea and anguish videos, then they answered their nausea and anguish perception intensity and unpleasantness using VAS.

#### Step 4 (Association between trait interoception and state interoception)

The correlation analysis was conducted between responses to the embodied interoceptive trait proto-questionnaire (step 2) and the perception and unpleasantness intensities reported during the interoceptive state test of each channel (step 3) using Pearson’s correlation.

#### Step 5 (Psychometric analyses)

##### Exploratory factor analysis

First, correlation matrices and plots were used as exploratory analysis tools to better understand the association across all items. Since items in our questionnaire are numeric, Pearson’s correlation tests were used. A series of exploratory factor analyses was also conducted using oblique and orthogonal rotations to explore different factorial solutions underlying the data, using maximum likelihood as the extraction method. Our heuristic for selecting factor solutions included: (a) scree plots, (b) solutions that were theoretically justifiable, and (c) solutions where items loaded with values above 0.30 on a single factor while all other loadings were below that threshold.

##### Confirmatory factor analysis

A series of confirmatory factor analyses (CFA) was then used to evaluate the latent variables. Our model was theoretically justified fitting statistics for confirmatory factor analyses, which included Chi-squared value based on the fit function, Degrees of freedom for model, P-value for chi-squared value and degrees of freedom, Comparative Fit Index, Non-Normed Fit Index, Incremental Fit Index, Relative Noncentrality Index, Root Mean Square Error of Approximation, Root Mean Square Residual (includes means), Root Mean Square Residual (no means), Standardized Root Mean Square Residual (includes means), Bentler Standardized Root Mean Square Residual (includes means), Bentler Standardized Root Mean Square Residual (no means), MPlus Standardized Root Mean Square Residual (includes means), MPlus Standardized Root Mean Square Residual (no means), Goodness-of-Fit Index, as well as Adjusted Goodness-of-Fit Index.

### Latent variable modeling

#### Item reduction using a rating scale stepwise reduction procedure

This method systematically reduces the number of response categories or items while maintaining or improving the scale’s psychometric properties. The process continues until the scale achieves an optimal balance between simplicity and maintaining information integrity.(DeVellis & Thorpe, 2021) The goal is to eliminate redundant or ineffective categories that do not provide additional useful information, thus improving the reliability, validity, and interpretability of the scale. A shorter, more streamlined scale is easier for respondents to complete and less cognitively demanding.(Lozano et al., 2008)

#### Item response theory (IRT)

IRT modeling was conducted to identify the level of interoceptive experiences associated with each item (question). To accomplish this matching between items and interoceptive perception intensity, and unpleasantness level, graded models were used since all constructs presented ordinal item types. The graded response model(Samejima, 1969) is an IRT model developed to evaluate surveys with ordinal responses such as ordered Likert-type scales. This model characterizes item functioning via two parameters for each item: item discrimination and item difficulty. Discrimination parameters evaluate how well an item discriminates (or differentiates) between individuals scoring high and low on a construct. Item difficulty (or item location) describes how much of a certain trait or ability on a construct a subject typically possesses before they endorse a specific item on a test or questionnaire.(Depaoli et al., 2018; Hambleton et al., 1991) IRT modeling was evaluated using Item Characteristic Curves (ICC). The fit of our IRT models was evaluated using the root mean square error of approximation (RMSEA), with the criterion for acceptable fit being set at a value < 0.10.(Fieo et al., 2016)

#### Score development

The interoception trait score was calculated for each participant according to the following formula:

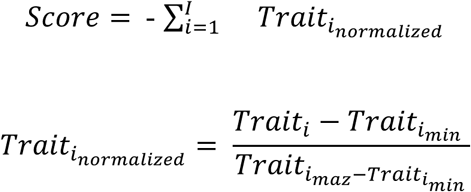

Where *i* represent each trait and *I* represent the total number of traits selected after item reduction.

#### Reliability analyses

##### Internal consistency

Guttman’s Lambda 6 and inter-item correlation under internal reliability metrics were reported.(Raykov & Marcoulides, 2011) Furthermore, internal consistency was analyzed using Cronbach’s alpha coefficient, the most widely used metric.(Streiner, 2003) Since raw Cronbach’s alpha can be sensitive to differences in the item variances, both raw and standardized alpha were reported, the latter being based on the correlations rather than the covariances. The minimum value considered acceptable for the Cronbach’s alpha coefficient was 0.7.(George & Mallery, 2003; Hopkins, 2000; Peterson & Kim, 2013) This evaluation reflects the extent to which questionnaire items are inter-correlated, or whether they are consistent in measurement of the same construct.

##### Test-retest reliability

Healthy volunteers were invited to answer the questionnaire 3 times for short- and long-term test-retest reliability. The interval between the first and second application of the instrument was three hours (‘short-term’ reliability) and the third was 30 days (‘long-term’ reliability). Test-retest reliability was measured with intraclass correlation coefficient (ICC) by using a two-way mixed model with absolute agreement. Based on the 95% confidence interval of the ICC estimate, reliability was considered low if < 0.5, moderate between 0.5 and 0.75, good between 0.75 and 0.9, and excellent if > 0.9.(Koo & Li, 2016)

#### Validity analyses

##### Convergent validity

Pearson’s correlation methods with heatmap plots was used to determine the association between embodied interoceptive questionnaire and previously established questionnaires that evaluate concepts related to interoceptive sensations in healthy volunteers: Multidimensional Assessment of Interoceptive Awareness (MAIA),(Mehling et al., 2012) Interoceptive Accuracy Scale (IAS),(Murphy et al., 2020) Interoceptive Confusion Questionnaire (ICQ),(Brewer et al., 2016; Goerlich, 2018) Body Perception Questionnaire (BPQ),(Cerritelli et al., 2021) and Toronto alexithymia scale (TAS-20).(Fortes et al., 2017)

##### Discriminant and divergent validities

In these validations, patients with neuropathic pain were evaluated. They answered the Intero-10, VAS, BPI,(Ferreira et al., 2011; Melzack, 1987) NPSI,(de Andrade et al., 2011) MOS,(Hays et al., 2005) HADS,(Zigmond & Snaith, 1983) and SF-6D.(Campolina et al., 2011) Discriminant validity was evaluated through inter-construct correlations between the Intero-10 questionnaire and NPSI using the Heterotrait–Monotrait Ratio (HTMT) method. HTMT calculates the mean correlation among indicators across different constructs (heterotrait– heteromethod correlations) relative to the mean correlation among indicators within the same construct (monotrait–heteromethod correlations). HTMT values below 0.90 indicate discriminant validity between two reflective constructs.(Franke & Sarstedt, 2019) To assess the divergent validity, Spearman’s rho was used to estimate the correlation between the Intero-10 and established chronic pain assessment tools.

##### Mediation analyses

Two models were used for mediation analysis. The first model assessed whether the association between pain interference and quality of life might be mediated by higher interoception scores, and the second model assessed whether the association between mood and quality of life might be mediated by higher interoception scores. These models were exploratory and informed by correlations identified in our previous aim and existing literature. Assumptions for linear regression analyses were met, and indirect, direct, and total effects were assessed to interpret mediation. Statistical significance was evaluated using 95% confidence intervals generated from 1,000 bootstrapped samples. Model fit was assessed by R^2^ values, and effect sizes assessed by bootstrapped β-coefficients.

##### Face and content validities in patients with chronic neuropathic pain

A random sample of potential users of Intero-10 was involved in this part of the study. They were recruited from pain clinics at Institute of Orthopedics and Traumatology of the Hospital das Clínicas, University of São Paulo. They were requested to evaluate each part of the tool (introduction, single question and items) with reference to its clarity and understandability.(Cocchi et al., 2023)

##### Other statisticmal methods

The R language was used for analyses. Our exploratory analysis started with a visual evaluation of all variables to assess the frequency, percentage, and near-zero variance for categorical variables (comorbidity, educational level, ethnicity, gender, and religion) and their corresponding missing value patterns.(Kuhn & Johnson, 2013) Near zero variance is found when a categorical variable has a small percentage of a given category and is addressed by combining different categories. For mediation analysis was used PROCESS (version 4.3.1) model 4 in R (version 4.4.0).(Hayes, 2022)

## Results

### Participants

In total 467 participants were included in this study, 381 healthy volunteers and 86 patients with neuropathic pain. Table 1 presents a sample description of volunteers. Health status was classified as good and very good in 92.2% of healthy volunteers and BMI (body mass index) was 25.5 ± 5.45.

**Table 1:** Sample characteristics.

| Variable | Healthy volunteers<br>(n = 381) | Patients with neuropathic pain<br>(n = 86) |
| --- | --- | --- |
| Age (years) | 29.10 ± 7.95 (18-56) | 51.22 ± 12.04 (23-85) |
| Female | 203 (53.3) | 35 (40.7) |
| Color/race |  |  |
| White | 200 (52.5) | 46 (53.5) |
| Black | 12 (3.1) | 12 (14) |
| Other | 79 (20.7) | 28 (32.6) |
| Not informed | 90 (23.6) | - |
| Education level |  |  |
| College degree | 93 (24.4) | 17 (19.8) |
| No college degree | 147 (38.5) | 69 (80.2) |
| Not informed | 141 (37.0) | - |
| Current occupation |  |  |
| Employed | 107 (28.1) | 23 (26.7) |
| Retired | 2 (0.5) | 41 (47.7) |
| Others | 180 (47.2) | 22 (25.6) |
| Not informed | 92 (24.1) | - |
| Religion |  |  |
| No | 4 (2.2) | 4 (4.7) |
| Yes | 107 (57.8) | 82 (95.3) |
| Not informed | 74 (40.0) | - |
| Marital status |  |  |
| Married | 57 (30.8) | 36 (41.9) |
| Other | 54 (29.2) | 50 (58.1) |
| Not informed | 74 (40.0) | - |
| Intero-10 (total score) | 4.93 ± 1.86 (0.19 - 9.80) | 4.31 ± 2.31 (0.50 - 9.60) |

Legend: Values are presented as mean ± standard deviation, or frequency (percentage).

### 3.2 Step 1 (Embodied interoceptive channel selection)

In total, 752 full-text articles were identified in the literature review, all of which had been published between 1976 and 2018. All articles not related to interoception, or bodily signs were excluded. After reviewing the available literature, 28 items related to internal sensations were identified as interoceptive channels: pain, heartbeat, respiratory frequency, thirst, hunger, heat, cold, itching, sleep, muscle fatigue, nausea, voices (in the case of schizophrenia), dizziness, tickling, numbness, sensation of muscle stiffness, tingling, stretching of the skin, electrical flow, needling, dyspnea, sexual arousal, anguish, and visceral sensations such as visceral pain, distention of the bladder, stomach, rectum, and esophagus.(A. D. Craig, 2002; A. D. (bud) Craig, 2003; Critchley & Harrison, 2013; DuBois et al., 2016; Khalsa et al., 2018; Khalsa & Lapidus, 2016) Then, embodied interoceptive channels were selected based on the relevance and feasibility of the respective experimental setup for each of them. The following channels were deemed relevant and feasible: pressure pain, visceral pain, heartbeat, respiratory frequency, hunger, thirst, heat, cold, itching, dyspnea, sleep, muscle fatigue, gastric fullness, nausea, and anguish. In total, the assessment of 15 different interoceptive channels was reproduced or adapted in a psychophysical setup in Step 3. For each of these interoceptive channels, each participant’s level of interoceptive perception intensity, and perception of unpleasantness were ranked in a 0-100 mm visual analogue scale (VAS).

### Step 2 (Embodied interoceptive trait proto-questionnaire)

#### Face and content validity

The total content validity coefficient (CVC*t*) values were deemed adequate, with all items scoring above 0.8 for both clarity and relevance, except for the first question (CVC*t* = 0.79). Supplementary Table S1 displays the CVC values for the 15 channels evaluated by the experts panel. They proposed two additional items—bladder fullness, and muscle soreness— due to their relevance on the field of interoception. This resulted in a total of 17 channels in the embodied interoceptive trait proto-questionnaire, with two questions per channel. Adjustments were made to the questions, when necessary, based on experts feedback, and agreement after rounds. Supplementary Text S2 shows the English final version of the embodied interoception trait proto-questionnaire.

#### Embodied interoceptive trait proto-questionnaire

In total, 381 healthy volunteers answered this trait proto-questionnaire (Supplementary Table S2).

### Step 3 (Interoceptive state assessment during psychophysics setups)

In this step, the interoceptive state perception intensity and unpleasantness levels in healthy volunteers were assessed and scored in a psychophysics setting for 16 embodied interoceptive channels (Fig. 2). Supplementary Table S2 presents the mean of the embodied interoceptive state assessment.

**Figure 2.**
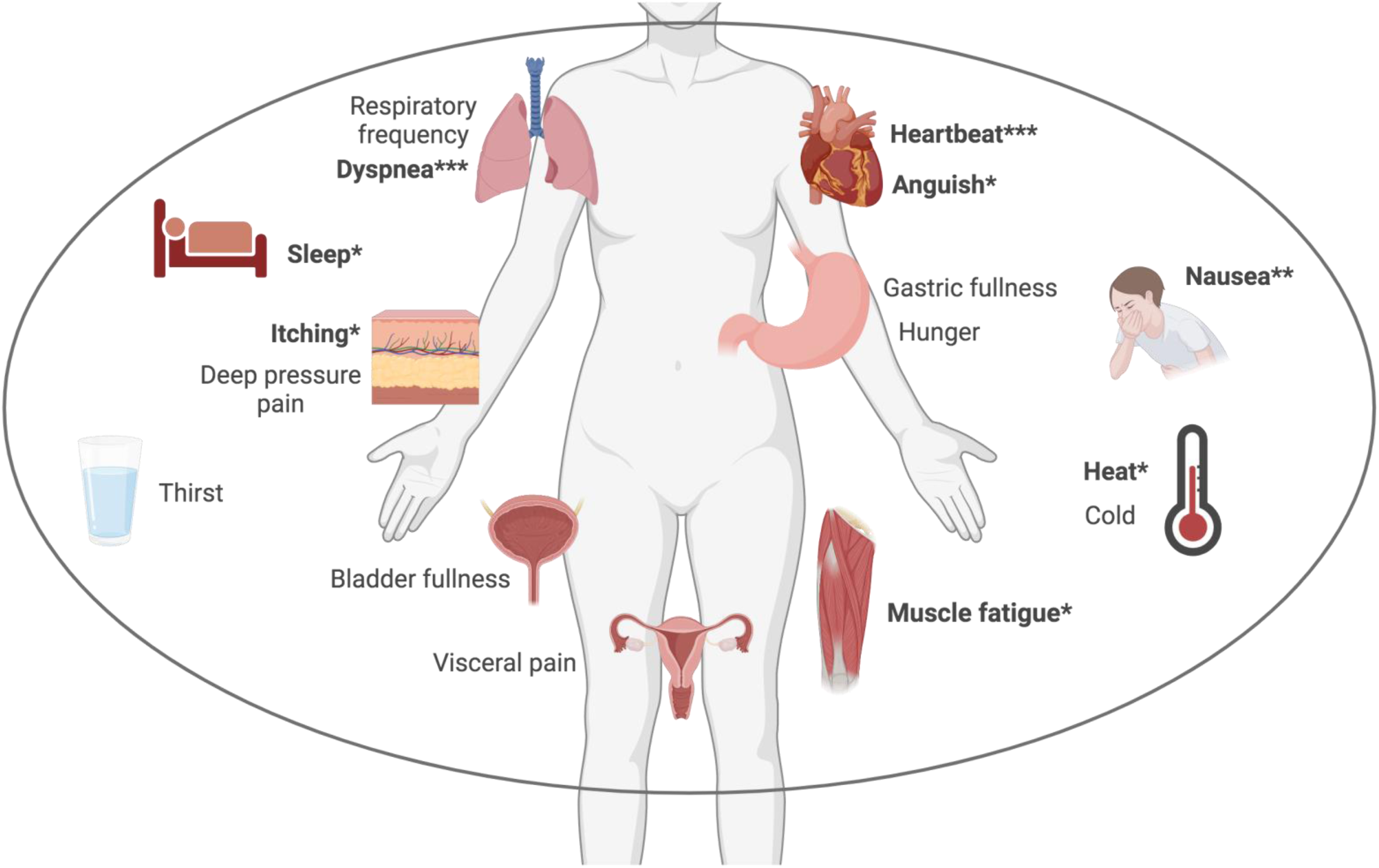
Embodied interoceptive channels. Sixteen embodied interoceptive channels assessed in step 3. *: channels included in the embodied perception intensity category of the Intero-10. **: channels included in the unpleasantness category of the Intero-10. ***: channels included in both categories (embodied perception intensity and unpleasantness).

### Step 4 (Association between trait interoception and state interoception)

Our results indicated a significant positive correlation between trait and state interoception scores for the following items: heartbeat unpleasantness (0.61, *P* = 0.001), respiratory frequency unpleasantness (0.47, *P* = 0.043), hunger unpleasantness (0.66, *P* = 0.004); cold unpleasantness (0.45; *P* = 0.048), heat unpleasantness (0.46, *P* = 0.030), anguish unpleasantness (0.42, *P* = 0.050), and gastric fullness unpleasantness (0.55, *P* = 0.015). A significant negative correlation was found in itching unpleasantness (-0.40, *P* = 0.050). The remaining interoceptive state vs. trait correlations were non-significant (*P* > 0 .050). For the sleep channel, the total sleep time (TST) and time in bed (TIB) self-reported were measured and correlated with TST and TIB real measured by actigraphy. The correlation for TST was 0.54, *P* = 0.010, and for TIB, it was 0.70, *P* = 0.001.

### Step 5 (Psychometric analyses)

#### Exploratory factor analysis

When evaluating the correlation matrix for the original 34 questions in the embodied interoceptive trait proto-questionnaire without transformation, no particular item concentration was observed, indicating a single factor for this scale. A series of exploratory factor analyses was then performed, including all 34 original items from the questionnaire. Factor analysis was run for the 34 original items, and identified a single factor structure, which was confirmed by plotting the factors (Supplementary Fig. S1).

#### Confirmatory factor analysis

Next, a confirmatory factor analysis was performed assuming a single factor for the embodied interoceptive trait proto-questionnaire. An overall good fit based on a wide range of metrics was indicated, including a root mean square error of approximation (RMSEA) of 0.092, a standardized root mean square residual (SRMR) of 0.095, Bentler SRMR (includes mean) of 0.092, Bentler SRMR (no mean) of 0.095, goodness-of-fit (GFI) index of 0.821, and adjusted goodness-of-fit (AGFI) index of 0.786. However, the metrics comparative fit index (CFI), tucker-lewis index (TLI), non-normed fit index (NNFI), normed fit index (NFI), incremental fit index (IFI), and relative fit index (RFI) were below the gold standard of good model fit (>0.95).(Depaoli et al., 2018; Samejima, 1969) Therefore, selected items were revised in our scale to improve the CFA fit.

#### Item reduction using a rating scale stepwise reduction procedure

Supplementary Table S3. presents the Area Under the Curve (AUC) for the 34 original items of the embodied interoceptive trait proto-questionnaire in the second column, sorted in ascending order. The variable X23 (nausea perception) was the item with the largest AUC. Supplementary Fig. S2 displays the stepwise reduction procedure for the questionnaire’s item reduction.(Koczkodaj & Wolny-Dominiak, 2017) The AUC of all 34 items of the trait interoception scale was presented. The AUC value increased by adding the first two items labeled X23 (nausea perception) and X22 (muscle soreness unpleasantness). The curve peeked at item X22. The addition of variable X19 (itching perception) resulted in a decrease in AUC. As a result, the reduction procedure was stopped after adding the first two items. This analysis assumes that selecting the item with the largest AUC can reduce the number of items. The main idea of this heuristic is to add the following question with the largest AUC to the selected item from the subset of the remaining items until the AUC of all items decreases.

Supplementary Fig. S3 displays Receiver Operator Characteristic (ROC curve of total AUC of items in reduced rating scale. The ROC indicates the overall accuracy and the separation performance of the rating scale without predictability loss.

#### Item response theory item reduction

A stepwise item reduction was performed based on the item information index obtained from the IRT analysis. In total, 24 items with a low information index were removed. The 10 items with greater discrimination were selected, as they are better able to detect subtle differences in the respondents’ abilities.(Baker & Kim, 2017; Embretson & Reise, 2013) The questionnaire after IRT item reduction included the following Supplementary Fig. S4 displays the scale information function for the unique construct before and after the item reduction process. Supplementary Table S4 shows IRT goodness-of-fit before and after item reduction. The final 10 items included heartbeat, heat, itching, dyspnea, sleep, muscle fatigue, and anguish for interoceptive perception intensity, while heartbeat, dyspnea, and nausea were selected for unpleasantness intensity (Fig. 2). Supplementary Fig. S5 shows the Intero-10 questionnaire (also available in three languages in a free app form https://neuro-science.vercel.app/.

#### Internal consistency

The internal consistency of Intero-10 was classified as adequate with a Cronbach’s alpha of 0.81 after reduction.

#### Score Development

The 10-item trait interoception score values ranged from 0 to 10. The calculation of normative data based on percentile allowed for the classification of any responderś scores as being low (0 - 4.25), medium (4.26 - 5.75), or high (5.77 - 10) scores based on data from healthy adults (Supplementary Table S5).

#### Convergent validity

A total of 102 healthy volunteers (68 female) between ages of 19 and 56 years old (M = 28.6, SD.= 7.39) participated in this validation. Participants completed the Intero-10 and other interoception-related questionnaires. Weak correlations were observed between the Intero-10 and the MAIA subscales: Body Listening (0.21; *P* = 0.030), Emotional Awareness (0.20; *P* = 0.030), Not-Worrying (0.25; *P* = 0.010), and Trusting (-0.26; *P* = 0.010). No significant correlations were found for the Noticing (0.07; *P* = 0.440), Not-Distracting (0.14; *P* = 0.140), Attention Regulation (0.05; *P* = 0.60), and Self-Regulation (0.02; *P* = 0.820) subscales. The Intero-10 showed moderate correlations with BPQ Awareness (0.42; *P* < 0.001) and BPQ Reactivity (0.50; *P* = 0.01). No significant correlations were observed between the Intero-10 and the IAS (0.11; *P* = 0.230), ICQ (0.02; *P* = 0.840), or TAS (0.17; *P* = 0.080) scales.

#### Test-retest reliability

The ICC average for the 10-item reduced scale was 0.86 (95% CI: 0.79-0.920), indicating good reliability. In the short-term analysis (n = 56), the ICC was 0.84 (95% CI: 0.75-0.90), while the long-term analysis (n = 44) yielded an ICC of 0.78 (95% CI: 0.66-0.87), both demonstrating good reliability. Supplementary Table S6 displays the ICC values for each item.

#### Discriminant validity

The inter-construct correlation (HTMT) between embodied interoception (Intero-10) and neuropathic pain symptoms (NPSI) was 0.74. Results under the cutoff of 0.9 means that discriminant validity has been established between variables.

#### Divergent validity

Intero-10 total score in patients with neuropathic pain was 4.31 ± 2.31 (0.50 - 9.60). Based on classification derived from data on healthy adults, 59% of patients were categorized as having low interoception (0 to 4.25), 15.1% as medium (4.26 to 5.76), and 25.6% as high (5.77 to 10).

Pain intensity was 79.65 ± 13.34 (40-100). SF-6D was 0.50 ± 0.21 (0-0.97), BPI severity and interference scores were 7.08 ± 1.32 (3.50-10.00) and 6.49 ± 2.12 (0.57-10.00), respectively. HADS anxiety, depression, and total score were 6.87 ± 4.70 (0-21), 6.09 ± 5.33 (0-19), and 12.96 ± 9.17 (0-40), respectively. MOS was 43.13 ± 16.34 (16.11-80).

Intero-10 showed a negative correlation with quality of life in patients with neuropathic pain (-0.46; *P* < 0.001), a positive correlation with negative mood (0.40; *P* < 0.001), and a positive correlation with pain interference (0.27; *P* = 0.005). No significant correlations were found between Intero-10 and pain intensity (VAS), pain severity (BPI), neuropathic pain symptoms (NPSI), and sleep (MOS).

#### Mediation analysis

The first model assessed whether the association between pain interference and quality of life was mediated by higher embodied interoception scores. This first model suggested significant total effect (β = -0.057 [-0.065, -0.031]), indicating that QoL was -0.057 points lower for every 1 point increase in pain interference. When controlling for interoception, there was still a direct effect (β = -0.048 [-0.065, -0.031]) of pain interference on QoL. Moreover, a significant indirect effect was found, with interoception partially mediating the correlation between higher pain interference and lower QoL (β = -0.0093 [-0.021, -0.0015]), indicating that not only patients with higher pain interference are likely to have lower levels of QoL, but this correlation is partially mediated by higher interoception scores (Fig. 3a). Participants with higher pain interference also had higher interoception scores (a-path: β = 0.33 [0.12, 0.56]), and higher interoception was also related to a lower QoL (b-path: β = -0.028 [-0.048, -0.0081]).

**Figure 3.**
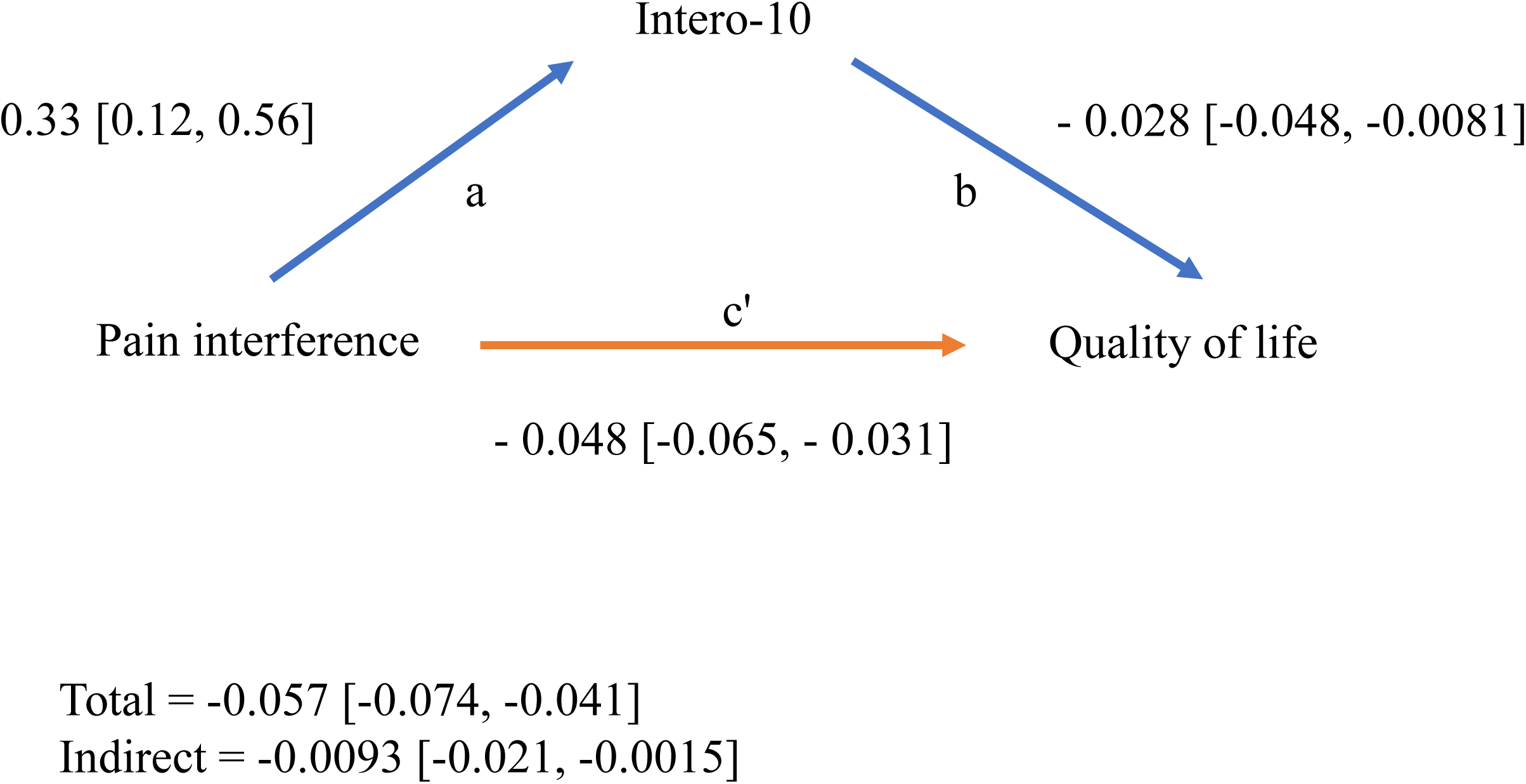
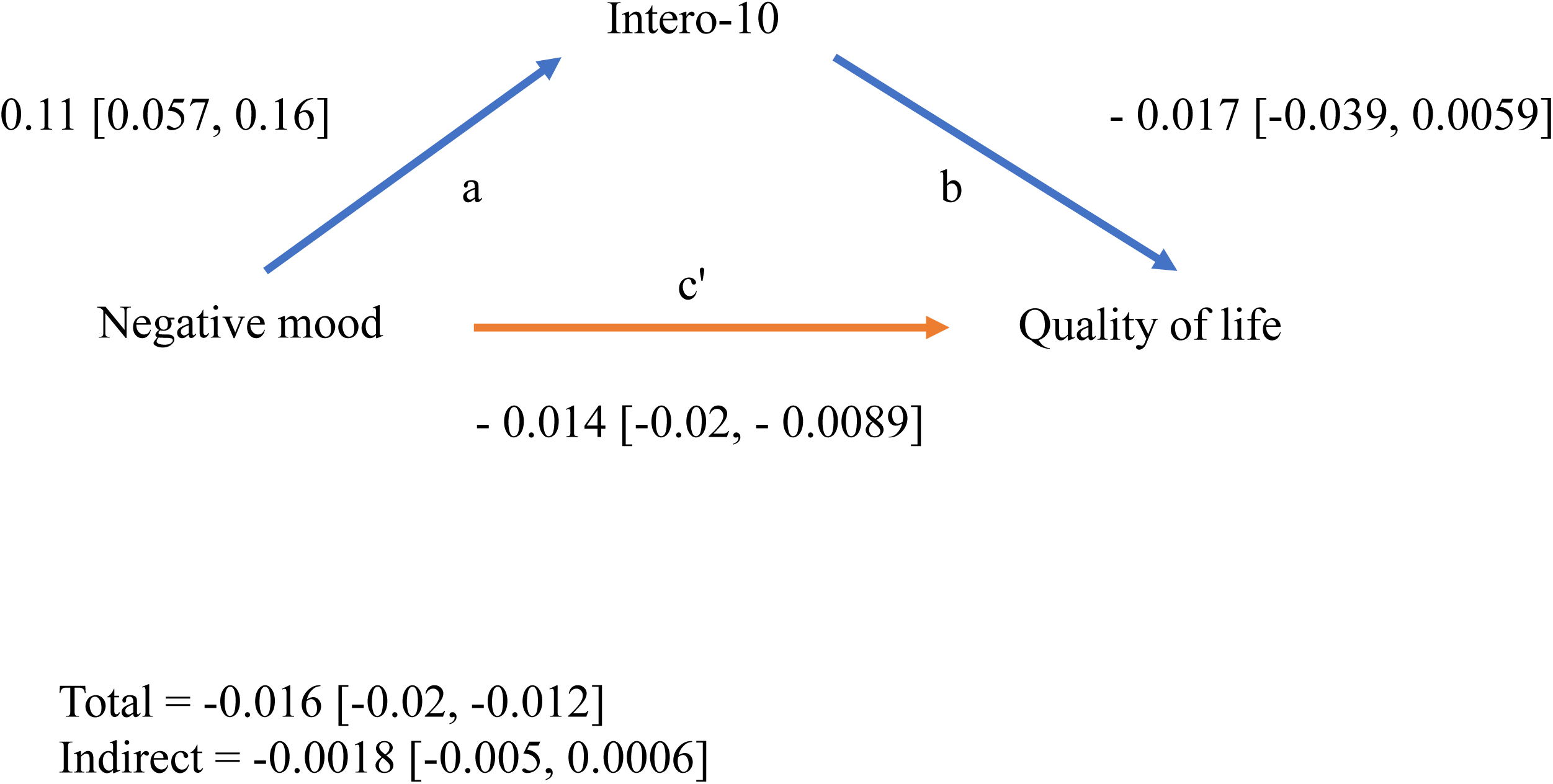
Mediation analyses. Figure 3a illustrates the mediation analysis model assessing the total and direct effects of pain interference on quality of life, with indirect effects via interoception (Intero-10). Blue arrows highlight the indirect effects (a and b-paths), and the orange arrow highlights the direct effect (c’-path). Figure 3b presentes the mediation analysis model assessing the total and direct effects of negative mood on quality of life, with indirect effects via interoception (Intero-10). Blue arrows highlight the indirect effects (a and b-paths), and the orange arrow highlights the direct effect (c’-path).

The second model assessed whether the association between negative mood and QoL might be mediated by higher interoception scores. This second model suggested a significant total effect (β = -0.016 [-0.02, -0.012]), indicating that QoL was 0.016 points lower for every 1 point increase in negative mood score. When controlling for interoception, there was also still a direct effect (β = -0.14 [-0.02,-0.0089]) of negative mood on QoL. However, no indirect effect was observed, as despite higher negative mood being related to higher interoception (a-path: β = 0.11 [0.057, 0.16]), higher interoception was not related to lower QoL (b-path: β = - 0.017 [-0.039, 0.0059]). Therefore, interoception did not mediate the relationship between mood and QoL (indirect effect: β = -0.0018 [-0.005, 0.0006]), demonstrating that patients with greater negative mood are likely to have lower levels of QoL, a relationship that is not explained by interoception levels (Fig. 3b).

#### Face and content validity in patients with chronic neuropathic pain

Five potential users (patients with chronic pain) were selected through a convenience sampling to evaluate the Intero-10 questionnaire in terms of clarity and relevance. Total CVC score (CVCt) for clarity was 0.9 and for relevance CVCt was 0.9. Supplementary Table S7 shows CVC for the 10 questions.

## Discussion

We report on the development and validation of a questionnaire to assess embodied interoception, based on the perceived intensity and unpleasantness of trait interoceptive channels. The Intero-10 relevant and discriminant psychometric items demonstrated adequate content validity, as assessed by experts and patients, good internal consistency, good reliability, and a single-factor structure. Additionally, it was reported which of the Intero-10 items correlated with “state” ratings of interoceptive experiences experimentally evoked during psychophysics setups. Finally, after calculation of normative data, the embodied interoceptive profiles of a sample of patients with neuropathic pain was reported, as well as correlations with other chronic pain domains and quality of life. These later analyses showed that higher embodied interoceptive scores correlated with worse mood symptoms and worse quality of life, and also partially mediated the correlation between pain interference and quality of life.

The choice of the potential embodied interoceptive items to be used in the Intero-10 was based on interoceptive channels previously reported in the literature as being testable by psychophysics setups. The original items included interoceptive perception intensity and unpleasantness scores of interoceptive channels (experiences) one routinely encounters in their daily lives. It has been shown that while bodily sensations can be perceived with varying degrees of intensity, their associated unpleasantness is an independent construct,(Rainville et al., 1997) which is believed to be processed by dedicated anatomical pathways, such as those having the anterior cingulate cortex as main hub.(A. D. B. Craig, 2003; Rainville et al., 1997) Such an approach is similar to the multidimensional characterization of pain, where both discriminative (intensity) and affective (unpleasantness) components are known to be relevant for pain description.(Melzack, 1987; Price et al., 1987) Participants “trait” scores based on the response to the questionnaire did not correlate to their experimentally evoked (state) scores during the lived experience of each of the interoceptive channels for intensity scores. Indeed, the relationship between self-reported and objective interoceptive measures has been investigated in several studies, with most studies pointing towards a weak correlation between self-report interoception and interoceptive accuracy (the degree of correspondence between actual and inferred bodily states).(Critchley et al., 2004; Ferentzi et al., 2018; Garfinkel et al., 2015; Mehling et al., 2012; Murphy et al., 2020) Different from intensity scores, unpleasantness ratings positively correlated between trait (questionnaire) and state (experimentally-evoked) embodied interoceptive channels. These included heartbeat, respiratory frequency, hunger, cold, heat, anguish, and gastric fullness. These original findings point towards a correlation between prediction and state-experience only for the unpleasantness, suggesting that the negative valence of internal experience provided a common currency for both state and trait ratings, with a potentially lower prediction error for unpleasantness when compared to intensity ratings of embodied interoceptive channels. The reasons for these findings need to be specifically addressed and could result from a stronger reliability of predictions for unpleasantness when compared to intensity ratings. Lower credibility for intensity ratings could lead to larger sensory modulation and gain in interoceptive inputs when estimating its intensity,(Seth, 2013) leading to potentially higher prediction error, to higher uncertainty and subsequently to lower correlation between trait prediction ratings and the assessments made when the embodied interoceptive experience is actually lived.

After item reduction, the most psychometrically relevant embodied interoceptive channels were included in the Intero-10. For embodied interoceptive intensity scoring, the most discriminant channels were heartbeat, heat, itching, dyspnea, sleep, muscle fatigue, and anguish. For embodied interoceptive unpleasantness, key channels were heartbeat, dyspnea, and nausea. Cardiorespiratory items, such as heartbeat and dyspnea, were present in both intensity and unpleasantness scores. Interestingly, these are equally the most studied interoceptive channels(Koreki et al., 2020; Seth et al., 2011) in general. Notably, cardiac interoception has been the focus of many studies, likely because it is easily measurable and quantifiable,(Critchley et al., 2004; Critchley & Garfinkel, 2017; Pollatos et al., 2005, 2007) and has been proposed to be a global marker of interoceptive accuracy in general, despite some controversies.(Desmedt & Van den Bergh, 2024)

The consolidated Intero-10 score ranged from 0 to 10. The calculation of normative data based on percentile allowed for the classification of responderś scores as being low (0 - 4.25), medium (4.26 - 5.75), or high (5.76 - 10), based on data from healthy adults. The measure of embodied interoception provides a complementary characterization to the already existing dimensions of interoception evaluated by former questionnaires, such as interoceptive awareness,(Mehling et al., 2012) interoceptive attention,(Gabriele et al., 2022) and interoceptive accuracy.(Murphy et al., 2020) The low convergence between the Intero-10 and other interoception questionnaires indicated that they assess different constructs, and are thus complementary and not interchangeable for the specific characterization of embodied interoceptive channels.(Desmedt et al., 2022; Vig et al., 2022) For instance, the Multidimensional Assessment of Interoceptive Awareness is a tool that evaluates various subscales, including emotional awareness and self-regulation,(Mehling et al., 2012) while the Body Perception Questionnaire measures body awareness and autonomic reactivity,(Cabrera et al., 2018; Cerritelli et al., 2021; Vig et al., 2022) the Interoceptive Accuracy Scale(Murphy et al., 2020) measures how accurately individuals perceive bodily sensations, and the Interoceptive Attention Scale(Gabriele et al., 2022) assesses the degree of focus on internal signals. The Intero-10 was not correlated with most of these scales, and only weakly or moderately correlated with certain domains of “body perception” (BPQ) and “body listening” (MAIA), for example. These results highlight that the Intero-10 may be used to assess specific constructs of interoception, mainly related to perceptions arising from body experiences.

Previous studies have shown that patients with chronic pain often exhibit low interoceptive accuracy, which has been negatively correlated with symptom’s severity in specific disorders.(Di Lernia et al., 2016) A recent meta-analysis evaluating the relationship between pain and interoception also indicated that individuals with chronic pain display increased interoceptive sensibility and reduced interoceptive accuracy.(Horsburgh et al., 2024) In the present study, while ca. 60% of patients with neuropathic pain were classified as having low embodied interoception scores, higher scores correlated with worse quality of life. Despite being related to different interoceptive domains, overestimations of predicted interoceptive perception have been reported in patients with functional neurological disorders, such as functional seizures, functional tremor, and movement disorders,(Apazoglou et al., 2017; Koreki et al., 2020; Pareés et al., 2012; Ricciardi et al., 2015) and is in line with reports of higher scores for domains such as interoceptive sensibility in people with chronic pain.(Horsburgh et al., 2024)

Attempts to frame and model chronic pain and some psychiatric disorders in terms of altered embodied interoceptive predictions have been put forth.(Critchley & Garfinkel, 2017; Di Lernia et al., 2016, 2020; K. Friston, 2010; K. J. Friston, 2017; Horsburgh et al., 2024; Sedley et al., 2024; Seth et al., 2011) Recent models propose for example that migraine may act as an allostatic reset triggered by unresolved interoceptive prediction errors. In this view, migraine serves as a ’failsafe’ mechanism, triggered by catastrophic prediction errors. When these errors cannot be corrected by adjusting physiological states or updating predictions, they are progressively amplified, leading to the premonitory phase of migraine and, eventually, to the characteristic intense symptoms of a migraine attack.(Sedley et al., 2024) The quantification of embodied interceptive experiences offer possibilities for research and therapeutic interventions on how maladaptive embodied interoceptive processes may contribute to various chronic pain syndromes and disease burden.

In the present study, high embodied interoception scores in patients with chronic neuropathic pain correlated with lower quality of life, increased pain interference, and negative mood. Furthermore, we found that embodied interoception mediates the relationship between pain interference and quality of life, which can offer mechanistic and therapeutic insights for future studies.(McCracken et al., 2022)

The present study has limitations, one of them being the possibility that relevant interoceptive channels were not included in the Intero-10 because they have not been previously reported to be evoked experimentally. Despite results on the role of embodied interoception quantification in people with neuropathic pain, people suffering from other chronic pain types were nor included here. It remains to be confirmed if the quantification of embodied interoceptive channels would be relevant in their clinical assessment. Another potential source of bias is that since neuropathic pain patients have a lesion or disease to the somatosensory system, lesions/diseases affecting interoceptive afferents could in theory influence the perception of interoceptive channels and influencing results *per se*, something that would not be expected in those not presenting with a disease that does not primarily affect the somatosensory system such as musculoskeletal pain. It is also unknown to which degree embodied interoceptive scores are sensitive to the effects of treatment. Our sample of patients were globally stable and medicated. One could hypothesize that embodied interoceptive scores may have been decreased by effective treatment in responders and remained high in those not experiencing satisfactory effects of therapy and thus remaining with low quality scores and high embodied interoceptive scores. This would explain the significant correlations between high Intero-10 scores, higher pain interference, and worse quality of life, while those with a satisfactory treatment response had Intero-10 scores ranked as low based on normative values from healthy people.

Indeed, several aspects of embodied interoception are currently unanswered, such as whether self-reported embodied interoception ratings change over time in disease, and whether their modulation would be a prognostic or therapeutic marker of clinical pain. This hypothesis can now be readily tested, explored and falsified with the Intero-10. We hope it will open new windows of insights,(Kiverstein et al., 2022; Sedley et al., 2024) and possibly foster new frameworks on how to explore the main factors driving pain development and chronification.

## Acknowledgements

The authors would like to thank the Pain Center - University of São Paulo, CAPES, and CAPES-PrInt for supporting this study, and to collaborators (Interoception Study Group), volunteers, and patients.

## Author contributions

AMF and DCA were collectively responsible for the conceptualization and design of the study, as well as the acquisition, analysis, and interpretation of data. They also played a crucial role in drafting the manuscript. DCA, MJT, and LTY were responsible for funding the study by the department’s pain center. DCA was the study supervisor. AFB was the study co supervisor, helping in part of data collection, discussion and revision of the manuscript. Interoception Study Group provided the study in different departments at the University of São Paulo. SKM was involved with data analysis, interpretation, and manuscript revision. All authors participated in discussions regarding the results, and they contributed to the revision and commentary process of the manuscript. The final version of the manuscript has received approval from all authors.

## Research funding

This study was funded by the Pain Center/HC-FMUSP, CAPES and CAPES-PrInt (Institucional Program of Internationalization). The Center for Neuroplasticity and Pain (CNAP) is supported by the Danish National Research Foundation (DNRF121). DCA is supported by a Novo Nordisk Grant NNF21OC0072828. AFB receives a scholarship for scientific productivity from CNPq.

## Data availability

The data supporting the main findings of this study can be made available to qualified investigators upon request to the corresponding author.

