## Supplemental material for "Embodied Interoception Questionnaire (Intero-10): development, validation, and application in people with chronic pain"

**Supplementary material**

**Supplementary Text S1.** Description of psychophysics setup in step 3.

i. Deep pressure pain perception: the FDX® algometer (Wagner instrument, Greenwich, USA) was used to induce deep pressure pain hyperalgesia. The algometer, with a 1.0 cm² rubber disk, was applied perpendicularly to the body surface at 1 kg/sec. The equipment registers, using a dynamometer, the pressure exerted on the compressed tissues, recorded in Kg/cm^2^. The procedure was conducted in a sound-attenuated, climate-controlled room, with participants seated comfortably. Before testing, volunteers were familiarized with the procedure. They were instructed to identify the pressure pain threshold (PPT). In this study, deep pressure was applied on the thenar eminence of the right hand. It was emphasized that the participant should immediately communicate the onset of any local painful sensation, and not tolerate pain. Following this, supra threshold pain stimulation was performed at 1.3 x PPT, with deep pressure applied for 5 seconds, following the protocol described previously [[23]](https://paperpile.com/c/po7QKR/yx2XJ). After each stimulus, participants rated pain intensity and unpleasantness using Visual Analogue Scale (VAS). This procedure was repeated twice for consistency.

ii. Visceral pain perception: healthy female volunteers undergoing routine Intrauterine Device (IUD) insertion were assessed using a standard procedure [[18]](https://paperpile.com/c/po7QKR/C33n). The intensity of visceral pain experienced during cervical clamping was measured in seconds.  The procedure was performed by a gynecologist who noted the start (cervical gripping and traction) and end of the insertion. Volunteers were informed that an evaluator would observe without interfering. After completing the procedure, participants assessed the visceral pain perception and unpleasantness using VAS. To avoid bias related to technique or procedure time, only one gynecologist performed the insertions. The time interval of visceral pain caused by cervical pressure during the IUD insertion was used as the quantitative measure.

iii. Heartbeat perception: To assess heartbeat perception, the Heartbeat Detection Tracking Task was employed, following Schandry's method [[25]](https://paperpile.com/c/po7QKR/E1x2G). Healthy participants lay supine, relaxed for 30 seconds, and had their baseline heart rate recorded for 3 minutes. They were instructed to mentally count their heartbeats without touching their wrist or using any other physical manipulation to aid heartbeat detection. The task was performed in three intervals of 25, 35 and 45 seconds each, each followed by a 30-second rest. Acoustic signals indicated the start and end of each random phase. After the sign-off, participants were asked to report the number of heartbeats counted. None of them were aware of the duration of the tasks or their performance. Heart rate was monitored using a Polar® RS800CX device, which transmitted the heartbeat data to the Kubios software for interpretation. In this device, a strap with electrodes, placed on the chest, captures the electrical impulses from the heart and transmits them through an electromagnetic field to the monitor. The captured signal is sent via an interface (via infrared) to the Polar ProTrainer5^TM^ software. Heartbeat data were transmitted to the Kubios program [[27]](https://paperpile.com/c/po7QKR/0P1hO) to be archived and interpreted. This HR monitor is a cost-effective, practical tool with a high correlation [[10,16]](https://paperpile.com/c/po7QKR/m7mq7+sP9dP). After this protocol, volunteers were asked how well they thought they counted their heartbeat and how unpleasant they felt during the task. This assessment is related to interoceptive awareness, an aspect of interoception assessed via metacognitive judgments of the interoceptive accuracy [[11]](https://paperpile.com/c/po7QKR/iyaha).

iv. Hunger perception: inclusion criteria for this step were: healthy and eutrophic volunteers (body mass index-BMI between 18.5 and 24.9 kg/m^2^), fasting for at least 12 hours and no eating disorders. Participants rated their hunger perception intensity and unpleasantness after fasting, and after ingesting a meal test. The biological measure regarding the interoceptive perception of hunger was the dosage of the ghrelin and leptin hormones, which were collected from the blood plasma. This dosage was collected after 12 hours fasting, 60 minutes, and 120 minutes after consuming a normocaloric, normolipidic and normoprotein liquid diet (Ensure^TM^, Abbott, EUA) [[20]](https://paperpile.com/c/po7QKR/A2Ax8). A purple tube – K2 EDTA (4 ml) – (BD® Ref: 367861) was used to collect the plasma together with a protease inhibitor (Roche Cocktail). The collected samples were centrifuged at 2,800 rpm at 4°C, aliquoted into 500µl microtubes and stored at -80°C until biochemical analysis following the protocol for using the Protease Inhibitor Cocktail (Complete^TM^ Mini, EDTA-free, Roche LifeScience, EUA). Ghrelin and leptin hormones analyses were performed with the Multiplex kit (LUMINEX – xMAP®)[[13]](https://paperpile.com/c/po7QKR/YX3sd). This technique consisted of the use of fluorescent-colored microspheres (probes) that covalently bound to capture antibodies. Capture antibodies were placed directly against the desired biomarker and, after a series of washes to remove unbound proteins, detection antibodies were added to create the sandwich complex. Final detection of the complex was formed by the addition of streptavidin-phycoerythrin conjugate, where phycoerythrin served as a fluorescent indicator. For this assay, 96-well plates with negative control and a customized panel with active ghrelin and leptin analytes (MILLIPLEX® Multiplex Assays Using Luminex®) were used, analyzed by the software is Bio-plex Manager Software (version 6.0).

v. Thirst perception: healthy and eutrophic volunteers consumed salty potato chips to induce thirst according to an adapted protocol [[6]](https://paperpile.com/c/po7QKR/yU2TX). Participants with pre-existing medical or psychological conditions that could affect weight-maintenance, diet, thirst or performance on tests, were excluded. Participants were instructed not to ingest any solid or liquid food for at least 2 hours before the test. They answered the thirst perception intensity and unpleasantness using VAS at 3 different moments. The first assessment was performed before starting the test (T1). Then, participants were asked to consume 19 grams of potato chips with 0.28 grams of salt in 10 minutes. After this period, volunteers answered the thirst perception and unpleasantness using VAS (T2) and they also answered it after an interval of 30 minutes (T3). Participants were provided with an opaque white plastic cup containing cold water, with the volume concealed. Subsequently, they were asked if they desired additional water. Participants had a brief break. After a 5-minute interval, they were asked if they would like more water and their cups were refilled upon request. The amount of water consumed was the quantitative measure of this assessment.

vi. Environmental temperature perception: healthy volunteers were exposed to rooms that could be hot (38°C on average) or cold (16°C on average), then we measured the ear temperature of the participants. These protocols were adapted from a previous study [[29]](https://paperpile.com/c/po7QKR/3Pujy). The air temperature and the relative humidity (RH) were controlled, which were measured and recorded by a digital thermo-hygrometer (Model 7666.02.0.00, INCOTERM®). The effective temperature (ET) was calculated by the equation proposed by Missenard (1933) [[22]](https://paperpile.com/c/po7QKR/jSl2y):

ET = air temperature – 0.4 x (1 – 0.01RH) x air temperature – 10

a) heat perception: first, participants were seated at rest for 20 minutes before the test in the waiting room at a comfortable temperature of about 24°C. They wore standard clothing provided. Then, they were taken to the test room (heated environment) where they were seated for 20 minutes at a temperature on average 38°C. They evaluated perception and unpleasantness by VAS before and after they were exposed to the heated environment. Ear temperature was measured with a digital thermometer by infrared sensor (INCOMED®) immediately after resting for 20 minutes and after 20 minutes of the heat test phase.

b) cold perception: the evaluation of thermal perception to cold followed the same heat protocol. However, after volunteers were at rest for 20 minutes, they were taken to a refrigerated environment at an average temperature of 16°C. Participants wore standard clothing for the test and were exposed to the cold for 20 minutes.

vii. Skin itching perception: skin prick testing (SPT) with histamine dihydrochloride (10 mg/ml in 50% glycerin solution with 0.4% phenol) was used to induce itching according to adapted protocol [[12]](https://paperpile.com/c/po7QKR/Imrb7). The exclusion criteria were: history of psoriasis, neurodermatitis, eczema or other skin diseases in the test area, those who were taking antihistamines or other medications, such as antidepressants or calcineurin inhibitors, which could interfere with the proper test interpretation. Before the experiment, healthy volunteers remained comfortably seated for 5 minutes at rest. Then, their arm was propped up on a table and the skin in the test area was wiped with alcohol. An area of 25 cm^2^ in the left forearm (on the surface of the brachioradialis muscle, anterior part) was marked. A single-headed metal lancet was pressed through a drop of histamine perpendicularly to the predetermined area of skin for a second. All tests were performed by the same trained investigator to minimize variability in the application technique. Participants evaluated the itching perception intensity and unpleasantness using VAS after 1, 5, and 10 minutes of the histamine administration. Then, the papule diameter and the area of hyperemia were calculated. These were quantitative measures used in this assessment.

viii.  Dyspnea perception: healthy volunteers underwent an inspiratory endurance test to assess their dyspnea perception intensity and unpleasantness using a PowerBreathe (Medical KH2®, IMT Technologies Ltd, Birmingham, UK). Maximal inspiratory pressure was assessed using the sustained maximum inspiratory pressure (SMIP) method. After 10 minutes of rest, participants performed the test following recommendations previously demonstrated [[7]](https://paperpile.com/c/po7QKR/g1hlc). During the test, the nose clip was used to prevent air leakage, and participants were instructed to keep their lips sealed around the mouthpiece. Participants were familiarized with the equipment before the test. Peak pressure (PP), maximum inspiratory pressure (MIP), mean pressure (MP), area under the work curve of the sustained inspiratory musculature, maximum inspiration time, and slope of the inspiratory curve were variables obtained.

Endurance test: 60% load of the MIP for 7 minutes was used to assess the inspiratory resistance test as previously described [[7]](https://paperpile.com/c/po7QKR/g1hlc). After each minute, volunteers assessed their dyspnea perception intensity and unpleasantness using VAS. Variables used for a quantitative analysis included: target load (cmH_2_0), energy best inspiratory (Joules), total energy (Joules), average pressure (cmH_2_0), average power (Watts), average flow (L/s), and average volume (L).

ix. Sleep duration perception: healthy volunteers used a sleep self-reported questionnaire and an actigraph (Act Trust® da Condor Instruments, São Paulo, Brasil) to register sleep parameters commonly estimated in sleep logs. This device detects body movements through an accelerometer system, skin temperature (˚C), and light intensity. It is used to study the sleep-wake cycle and its circadian rhythms, making it possible to draw a graph of the periods of sleep and wakefulness [[2]](https://paperpile.com/c/po7QKR/F5fx4). The actigraph is low cost compared to polysomnography (considered the gold standard) and it has a reliability coefficient of 0.8 to 0.9 [[21,28]](https://paperpile.com/c/po7QKR/ApCBo+dZew8). The correlation between these two methods is 0.97 when assessing total sleep time [[14]](https://paperpile.com/c/po7QKR/bn3cL). Participants received instructions on how to use the actigraph and they were asked to wear it on the wrist of the non-dominant arm continuously for five days, except when showering or on any occasion that could damage the device. They were also asked to complete a sleep diary where they had to record information about the time they went to bed, sleep onset and offset times, possible naps and any wakefulness during the test period. This evaluation followed The American Academy of Sleep Medicine recommendations [[26]](https://paperpile.com/c/po7QKR/oEqFe). The aim of this assessment was to measure and correlate the self-reported total sleep time (TST) and time in bed (TIB) data with these data obtained from actigraphy.

x. Muscle fatigue perception: This study took place at the Sports Medicine Department of University of São Paulo. Healthy volunteers underwent exercise testing on an ergometric bicycle (bike, Cybex®, USA) using the modified Astrand protocol to assess muscle fatigue [[4]](https://paperpile.com/c/po7QKR/2wzi9). This test recommends a speed of 60 revolutions per minute (rpm) with progressively increasing loads. Each stage of the test lasted 2 minutes and the load was increased by 25 watts every two minutes continuously until reaching maximum effort. The maximal effort test was ended by exhaustion or when the individual could no longer maintain the pre-established speed of 60 rpm. During the test, the volume of oxygen consumed (VO2) per minute, the production of carbon dioxide (VCO2) per minute, and the respiratory exchange ratio (RER) were recorded by ergospirometry (exhaled gas analysis) using an analyzer computerized gas metabolic rate (CPX/Ultima, MedGraphics®, St. Paul, MN, USA) [[30]](https://paperpile.com/c/po7QKR/5IAYo). In addition, the electrocardiogram (ECG) was recorded at rest, during exercise and in the recovery phase, using computerized ECG with 12 simultaneous leads (HeartWare®, Brazil) to verify the heart rate (HR) response. Systolic (SBP) and diastolic (DBP) blood pressure were also determined using an aneroid sphygmomanometer (Tycos®). The subjective perception of exertion was quantified by the Borg scale from 6 to 20 [[5]](https://paperpile.com/c/po7QKR/srEDE). The muscle fatigue perception intensity and unpleasantness were assessed by VAS after each 2-minute interval. Each volunteer was instructed to assess fatigue, especially in the quadriceps muscle. Two trained evaluators conducted the experiment. For the analysis of the quantitative variables, respiratory quotient (RQ) measures, VO2 (ml/kg/min), metabolic efficiency (ΔVO2/ΔWatts), oxygen pulse (mL) and HR were used in four moments: at the beginning of the test, at ventilatory thresholds 1 and 2 (VT1 and VT2), and at the end of the test. Furthermore, also in these four moments, muscle fatigue perception intensity and unpleasantness were assessed.

xi. Bladder fullness perception: healthy volunteers with IPSS (International prostate symptom score) < 8 and quality of life related to urinary symptoms equal to excellent, well or satisfied were included in this evaluation. After spontaneous urination, participants were asked to drink 500 mL of water every half hour and report when they reached three standard cystometric urgency thresholds: first sensation, first desire to urinate and maximum capacity [[1]](https://paperpile.com/c/po7QKR/cGNQS). At baseline voiding and at each of these time points, bladder volume was measured with three-dimensional ultrasonographic measurements, and participants assessed bladder fullness perception intensity and unpleasantness using VAS. The following text with instructions related to the times points was made available to the participants:

"Hello! Thank you very much for your participation. Please, let us know these three moments:

1) I feel like I already have some urine in my bladder, but I do not feel like urinating;

2) I feel a weak urge to urinate, but I still do not want to urinate;

3) I feel a strong urge to urinate, I want to urinate now".

xii. Respiratory frequency and gastric fullness perception: healthy individuals with no history of gastric disorders, such as gastroparesis and functional dyspepsia were included in this assessment. Participants underwent the Electrogastrography (EGG) examination and a Walter Load Test (WLT) protocol [[9,15]](https://paperpile.com/c/po7QKR/hcVzq+lpps6) for induction of gastric fullness. EGG is a noninvasive technique used to assess the myoelectrical activity of the stomach through electrodes placed on the abdominal surface [[19]](https://paperpile.com/c/po7QKR/4sgu6). Multichannel EGG was performed with the Polygraf Solar EGG module electrogastrography system developed by Medical Measurement Systems – MMS (Laborie Europe, Enschede, The Netherlands), which is composed of six bipolar electrodes, one reference electrode, one ground electrode, and a strap to record respiratory movement [[8]](https://paperpile.com/c/po7QKR/I153i). The skin preparation, electrode placement, and subject position were adapted from previous protocol [[8]](https://paperpile.com/c/po7QKR/I153i). During 10 minutes the EGG tracing was stabilized without recording. Then, the respiratory strap placed in the lower thoracic region registered the respiratory frequency during the next 30 minutes while the participant performed the respiratory frequency mental tracking task. This task was the same as the protocol used in evaluating heartbeat perception [[24,25]](https://paperpile.com/c/po7QKR/E1x2G+LgDGo), however participants were asked to mentally count their respiratory frequency. Afterwards, just as heart rate evaluation, the evaluator asked how well they thought they counted their respiratory frequency and how unpleasant they felt during the task. This evaluation also is related to metacognitive awareness of interoceptive sensitivity. Subsequently, the volunteers underwent the WLT with water ingestion in a period of 5 minutes until reaching the point of perceived gastric fullness [[9,15,17]](https://paperpile.com/c/po7QKR/lpps6+hcVzq+Xzzew). Participants were instructed to stop drinking when they felt completely full. The water was consumed from an unmarked bottle that would be refilled after each drink. The total volume consumed was calculated at the end of the task. Participants were blinded to the actual volume of water consumed. Finally, volunteers assessed gastric fullness perception intensity and unpleasantness using VAS. EGG recordings were taken during the WLT and continued for another 30 minutes [[9,15]](https://paperpile.com/c/po7QKR/hcVzq+lpps6).

xiii. Nausea and anguish perception: A specialist committee experienced in validating a set of emotional facial expressions selected a 1-minute-and-40-second video from a group of videos that could evoke a sensation of nausea. Additionally, the committee selected a 2-minute-and-30-second video that could provoke a sensation of anguish. Volunteers watched nausea and anguish videos, then they answered their nausea and anguish perception intensities and unpleasantness levels using VAS.

**Supplementary Table S1.** Content validity coefficient for the introduction and 15 items originally accessed by experts.

|  | **Clarity** | | | **Relevance** | | |
| --- | --- | --- | --- | --- | --- | --- |
|  | **Mean** | **CVC*i*** | **CVC*t*** | **Mean** | **CVC*i*** | **CVC*t*** |
| Introduction | 3.6 | 0.90 | 0.89 | 4.0 | 1.00 | 0.99 |
| Deep pressure pain | 3.2 | 0.80 | 0.79 | 3.2 | 0.80 | 0.79 |
| Visceral pain | 3.8 | 0.95 | 0.94 | 3.8 | 0.95 | 0.94 |
| Heartbeat | 4.0 | 1.00 | 0.99 | 4.0 | 1.00 | 0.99 |
| Respiratory frequency | 4.0 | 1.00 | 0.99 | 4.0 | 1.00 | 0.99 |
| Hunger | 3.4 | 0.85 | 0.84 | 3.8 | 0.95 | 0.94 |
| Thirst | 3.8 | 0.95 | 0.94 | 3.8 | 0.95 | 0.94 |
| Cold | 3.8 | 0.95 | 0.94 | 3.8 | 0.95 | 0.94 |
| Heat | 3.6 | 0.90 | 0.89 | 4.0 | 1.00 | 0.99 |
| Itch | 3.8 | 0.95 | 0.94 | 3.8 | 0.95 | 0.94 |
| Dyspnea | 3.8 | 0.95 | 0.94 | 3.8 | 0.95 | 0.94 |
| Nausea | 3.8 | 0.95 | 0.94 | 3.8 | 0.95 | 0.94 |
| Sleep* | 3.8 | 0.95 | 0.94 | 3.8 | 0.95 | 0.94 |
| Muscle fatigue | 3.8 | 0.95 | 0.94 | 3.8 | 0.95 | 0.94 |
| Anguish | 3.8 | 0.95 | 0.94 | 3.8 | 0.95 | 0.94 |
| Gastric fullness | 3.8 | 0.95 | 0.94 | 3.6 | 0.90 | 0.89 |

Legend: CVC*i* : initial content validity coefficient, CVC*t*: total content validity coefficient

**Supplementary Text S2.** Embodied interoceptive trait proto-questionnaire.

**Embodied interoceptive trait proto-questionnaire**

**Introduction:** We can all perceive different sensations in our everyday lives, like hot, cold, pain, hunger or thirst. These sensations are personal and vary from one person to another. We want to know to what extent you perceive these sensations from inside of your body and / or to what extent some of them may bother you when they are present. For each question, you will see a line that indicates the increase of intensity from the left (lower intensity) to the right (higher intensity). The measuring scale ranges from "I do not perceive it at all / I am not bothered at all" on the leftmost position, to "I perceive it at the maximum possible intensity / It bothers at the maximum possible intensity" in the rightmost position. Please mark on the line what the intensity is for each question.

1) To what extent do you perceive the feeling of pressure pain (caused by contact with other people or to objects, e.g., bumps, tightness of shoes, clothes, backpacks or purses) in your daily life?

I do not perceive it at all____________________________________________________ I perceive it at the maximum possible intensity

1.1) To what extent does it bother you to feel pressure pain in your daily life?

I am not bothered at all____________________________________________________ It bothers at the maximum possible intensity

2) To what extent do you perceive the feeling of cramps (intestinal, menstrual) or stomach pain in your daily life?

I do not perceive it at all____________________________________________________ I perceive it at the maximum possible intensity

2.1) To what extent do it bother you to feel cramps or stomach pain in your daily life?

I am not bothered at all____________________________________________________ It bothers at the maximum possible intensity

3) To what extent do you perceive your heart/chest palpitations in your daily life?

I do not perceive it at all____________________________________________________ I perceive it at the maximum possible intensity

3.1) To what extent does it bother you to feel heart/chest palpitations in your daily life?

I am not bothered at all____________________________________________________ It bothers at the maximum possible intensity

4) To what extent do you perceive your breathing in your daily life?

I do not perceive it at all____________________________________________________ I perceive it at the maximum possible intensity

4.1) To what extent does it bother you to feel your breathing in your daily life?

I am not bothered at all____________________________________________________ It bothers at the maximum possible intensity

5) To what extent do you perceive the feeling of hunger in your daily life?

I do not perceive it at all____________________________________________________ I perceive it at the maximum possible intensity

5.1) To what extent does it bother you to feel hungry in your daily life?

I am not bothered at all____________________________________________________ It bothers at the maximum possible intensity

6) To what extent do you perceive the feeling of thirst in your daily life?

I do not perceive it at all____________________________________________________ I perceive it at the maximum possible intensity

6.1) To what extent does it bother you to feel thirsty in your daily life?

I am not bothered at all____________________________________________________ It bothers at the maximum possible intensity

7) To what extent do you perceive the feeling of being cold in your daily life?

I do not perceive it at all____________________________________________________ I perceive it at the maximum possible intensity

7.1) To what extent does it bother you to feel being cold in your daily life?

I am not bothered at all____________________________________________________ It bothers at the maximum possible intensity

8) To what extent do you perceive the feeling of being hot in your daily life?

I do not perceive it at all____________________________________________________ I perceive it at the maximum possible intensity

8.1) To what extent does it bother you to feel being hot in your daily life?

I am not bothered at all____________________________________________________ It bothers at the maximum possible intensity

9) To what extent do you perceive the feeling of itch in your daily life?

I do not perceive it at all____________________________________________________ I perceive it at the maximum possible intensity

9.1) To what extent does it bother you to feel itch in your daily life?

I am not bothered at all____________________________________________________ It bothers at the maximum possible intensity

10) To what extent do you perceive the feeling of breathlessness in your daily life?

I do not perceive it at all____________________________________________________ I perceive it at the maximum possible intensity

10.1) To what extent does it bother you to feel breathless in your daily life?

I am not bothered at all____________________________________________________ It bothers at the maximum possible intensity

11) To what extent do you perceive the feeling of nausea in your daily life?

I do not perceive it at all____________________________________________________ I perceive it at the maximum possible intensity

11.1) To what extent does it bother you to feel nauseous in your daily life?

I am not bothered at all____________________________________________________ It bothers at the maximum possible intensity

12) To what extent do you perceive whether you have slept well or not in your daily life?

I do not perceive it at all____________________________________________________ I perceive it at the maximum possible intensity

12.1) To what extent does it bother you to feel that you have not slept well in your daily life?

I am not bothered at all____________________________________________________ It bothers at the maximum possible intensity

13) To what extent do you perceive the feeling of muscle fatigue in your daily life?

I do not perceive it at all____________________________________________________ I perceive it at the maximum possible intensity

13.1) To what extent does it bother you to feel muscle fatigue in your daily life?

I am not bothered at all____________________________________________________ It bothers at the maximum possible intensity

14) To what extent do you perceive the feeling of anguish in your daily life?

I do not perceive it at all____________________________________________________ I perceive it at the maximum possible intensity

14.1) To what extent does it bother you to feel anguish in your daily life?

I am not bothered at all____________________________________________________ It bothers at the maximum possible intensity

15) To what extent do you perceive that you are full or satiated after eating or drinking enough in your daily life?

I do not perceive it at all____________________________________________________ I perceive it at the maximum possible intensity

15.1) To what extent does it bother you to feel full or satiated after eating or drinking enough in your daily life??

I am not bothered at all____________________________________________________ It bothers at the maximum possible intensity

16) To what extent do you perceive the need to urinate in your daily life?

I do not perceive it at all____________________________________________________ I perceive it at the maximum possible intensity

16.1) To what extent does it bother you to feel the need to urinate in your daily life?

I am not bothered at all____________________________________________________ It bothers at the maximum possible intensity

17) To what extent do you perceive muscle soreness in your daily life?

I do not perceive it at all____________________________________________________ I perceive it at the maximum possible intensity

17.1) To what extent does it bother you to feel muscle soreness in your daily life?

I am not bothered at all____________________________________________________ It bothers at the maximum possible intensity

**Supplementary Table S2.** Interoceptive trait and state assessments using visual analogue scale (0-100mm).

| **Interoceptive channels** | **Trait (n = 381)** | | **State** | | | |
| --- | --- | --- | --- | --- | --- | --- |
|  | **Perception (VAS)** | **Unpleasantness (VAS)** | **n** | **Perception (VAS)** | **Unpleasantness (VAS)** | **Missing** |
| Deep pressure pain | 51.38 ± 26.23 (0 - 100) | 38.74 ± 27.84 (0 - 100) | 29 | 41.84 ± 24.02 (4 - 88) | 34.10 ± 24.84 (0 - 84) | 0 |
| Visceral pain | 47.00 ± 30.14 (0 - 100) | 51.55 ± 30.37 (0 - 100) | 20 | 41.95 ± 28.95 (0 - 99) | 37.20 ± 28.61 (0 - 100) | 0 |
| Heartbeat | 33.06 ± 27.42 (0 - 100) | 29.60 ± 29.13 (0 - 100) | 28 | 47.75 ± 25.68 (2 - 80) | 21.25 ± 33.75 (0 - 91) | 0 |
| Respiratory frequency | 40.20 ± 27.19 (0 - 100) | 20.59 ± 23.73 (0 - 100) | 20 | 74.55 ± 26.42 (0 - 100) | 17.90 ± 27.07 (0 - 100) | 0 |
| Hunger | 63.05 ± 24.48 (0 - 100) | 60.40 ± 28.94 (0 - 100) | 20 | 47.80 ± 27.99 (5 - 99) | 17.90 ± 27.07 (0 - 100) | 0 |
| Thirst | 58.51 ± 25.92 (0 - 100) | 59.15 ± 30.29 (0 - 100) | 20 | 81.05 ± 22.30 (12 - 100) | 78.15 ± 27.71 (14 - 100) | 0 |
| Cold | 53.84 ± 27.47 (0 - 100) | 55.87 ± 30.20 (0 - 100) | 20 | 64.95 ± 25.50 (21 - 100) | 55.55 ± 34.60 (0 - 100) | 0 |
| Heat | 64.17 ± 26.18 (0 - 100) | 67.57 ± 28.75 (0 - 100) | 22 | 82.86 ± 19.99 (35 - 100) | 66.64 ± 25.71 (13 - 100) | 0 |
| Itch | 40.95 ± 29.10 (0 - 100) | 54.05 ± 30.39 (0 - 100) | 24 | 53.08 ± 30.69 (0 - 100) | 43.00 ± 31.33 (0 - 100) | 0 |
| Dyspnea | 27.32 ± 27.81 (0 - 100) | 53.25 ± 35.32 (0 - 100) | 23 | 65.47 ± 29.49 (10 - 100) | 65.95 ± 31.48 (6 - 100) | 0 |
| Nausea | 35.23 ± 28.88 (0 - 100) | 59.72 ± 32.05 (0 - 100) | 20 | 47.80 ± 39.75 (0 - 100) | 46.50 ± 39.60 (0 - 100) | 0 |
| Sleep* | 73.14 ± 24.60 (0 - 100) | 75.47 ± 24.84 (0 - 100) | 20 | - | - | 0 |
| Muscle fatigue | 65.45 ± 24.71 (0 - 100) | 69.16 ± 25.26 (0 - 100) | 19 | 50.47 ± 23.71 (20 - 92) | 37.78 ± 23.30 (0 - 87) | 1 |
| Anguish | 46.37 ± 29.98 (0 - 100) | 62.61 ± 29.99 (0 - 100) | 22 | 63.55 ± 28.80 (9 - 100) | 70.73 ± 34.14 (0 - 100) | 0 |
| Gastric fullness | 62.85 ± 25.63 (0 - 100) | 51.98 ± 30.24 (0 - 100) | 20 | 83.05 ± 13.30 (54 - 100) | 47.90 ± 26.86 (5 - 100) | 0 |
| Bladder fullness | 63.15 ± 24.34 (0 - 100) | 55.89 ± 29.62 (0 - 100) | 20 | 91.25 ± 9.94 (65 - 100) | 90.70 ± 12.55 (52 - 100) | 0 |
| Muscle soreness** | 50.62 ± 27.90 (0 - 100) | 52.50 ± 28.89 (0 - 100) | - | - | - |  |

Legend: Values are presented as mean ± standard deviation (minimum - maximum), VAS: visual analog scale, *For the sleep channel we measured the self-reported total sleep time (TST) and time in bed (TIB) during state assessment. **Muscle soreness channel was not evaluated during psychophysics setup.

**Supplementary Figure S1.** Scree plot in the exploratory factor analysis (EFA).

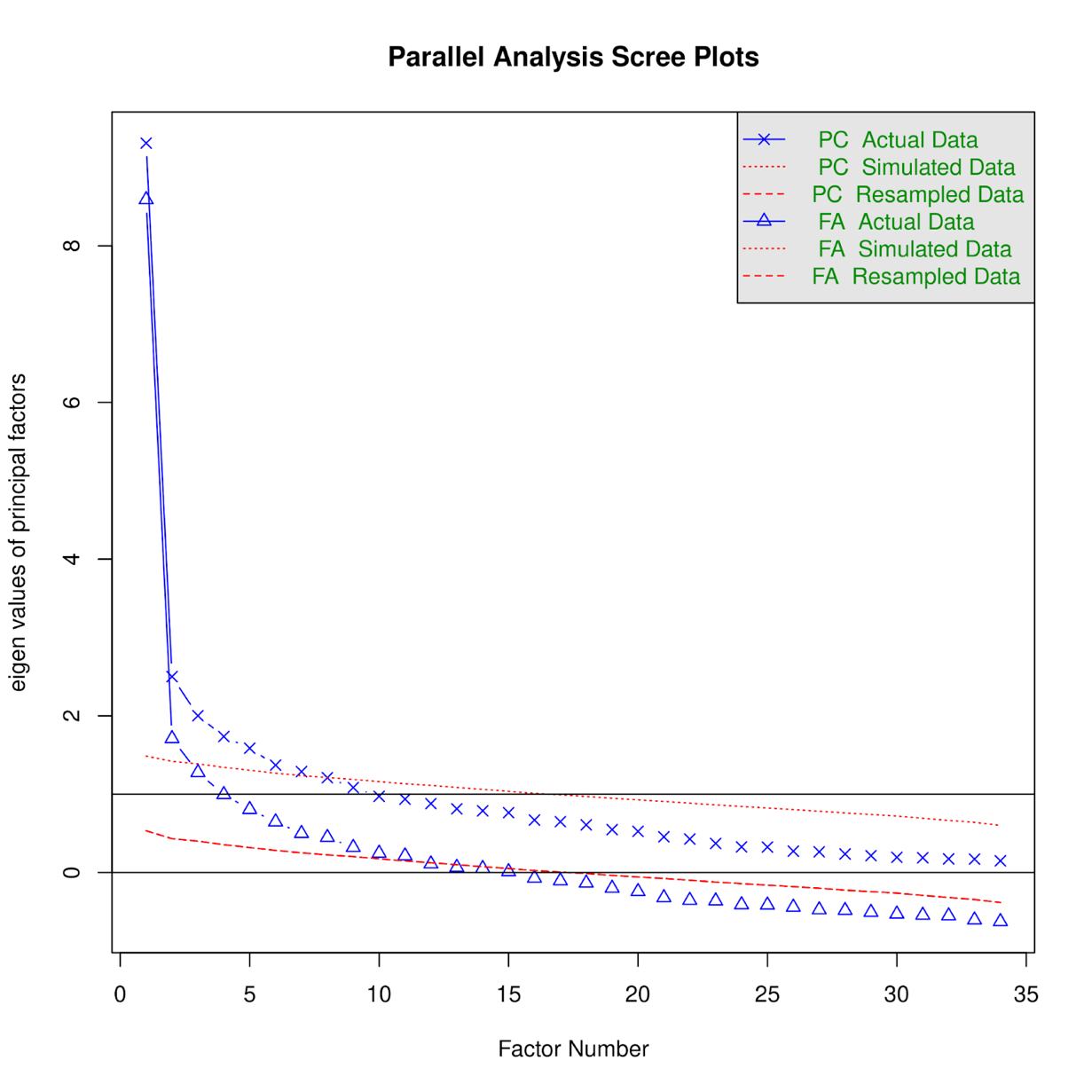

**Supplementary Table S3:** Stepwise AUC reduction from interoceptive trait scores.

| **Items** | | **AUC one variable** | **AUC running total** |
| --- | --- | --- | --- |
| X23 | (nausea perception) | 0.784 | 0.784 |
| X22 | (muscle soreness unpleasantness) | 0.780 | 0.803 |
| X19 | (itching perception) | 0.673 | 0.66 |
| X27 | (respiratory frequency perception) | 0.665 | 0.696 |
| X17 | (hunger perception) | 0.650 | 0.616 |
| X14 | (heat unpleasantness) | 0.635 | 0.553 |
| X25 | (deep pressure pain perception) | 0.618 | 0.508 |
| X33 | (visceral pain perception) | 0.616 | 0.526 |
| X26 | (deep pressure pain unpleasantness) | 0.610 | 0.491 |
| X5 | (cold perception) | 0.597 | 0.503 |
| X4 | (bladder fullness unpleasantness) | 0.592 | 0.516 |
| X12 | (gastric fullness unpleasantness) | 0.590 | 0.533 |
| X8 | (dyspnea unpleasantness) | 0.580 | 0.545 |
| X10 | (muscle fatigue unpleasantness) | 0.572 | 0.537 |
| X29 | (sleep perception) | 0.565 | 0.539 |
| X9 | (muscle fatigue perception) | 0.562 | 0.533 |
| X16 | (heartbeat unpleasantness) | 0.560 | 0.54 |
| X28 | (respiratory frequency unpleasantness) | 0.558 | 0.533 |
| X21 | (muscle soreness perception) | 0.556 | 0.538 |
| X34 | (visceral pain unpleasantness) | 0.555 | 0.54 |
| X6 | (cold unpleasantness) | 0.553 | 0.534 |
| X2 | (anguish unpleasantness) | 0.551 | 0.544 |
| X3 | (bladder fullness perception) | 0.547 | 0.548 |
| X25 | (hunger unpleasantness) | 0.539 | 0.546 |
| X31 | (thirst perception) | 0.534 | 0.545 |
| X1 | (anguish perception) | 0.531 | 0.541 |
| X15 | (heartbeat perception) | 0.529 | 0.535 |
| X11 | (gastric fullness perception) | 0.525 | 0.539 |
| X7 | (dyspnea perception) | 0.521 | 0.536 |
| X13 | (heat perception) | 0.519 | 0.535 |
| X20 | (itching unpleasantness) | 0.516 | 0.532 |
| X32 | (thirst unpleasantness) | 0.510 | 0.533 |
| X30 | (sleep unpleasantness) | 0.503 | 0.534 |
| X24 | (nausea unpleasantness) | 0.477 | 0.535 |

Legend: AUC: Area Under the Curve.

**Supplementary Figure S2:** Stepwise AUC reduction from interoceptive trait scores.

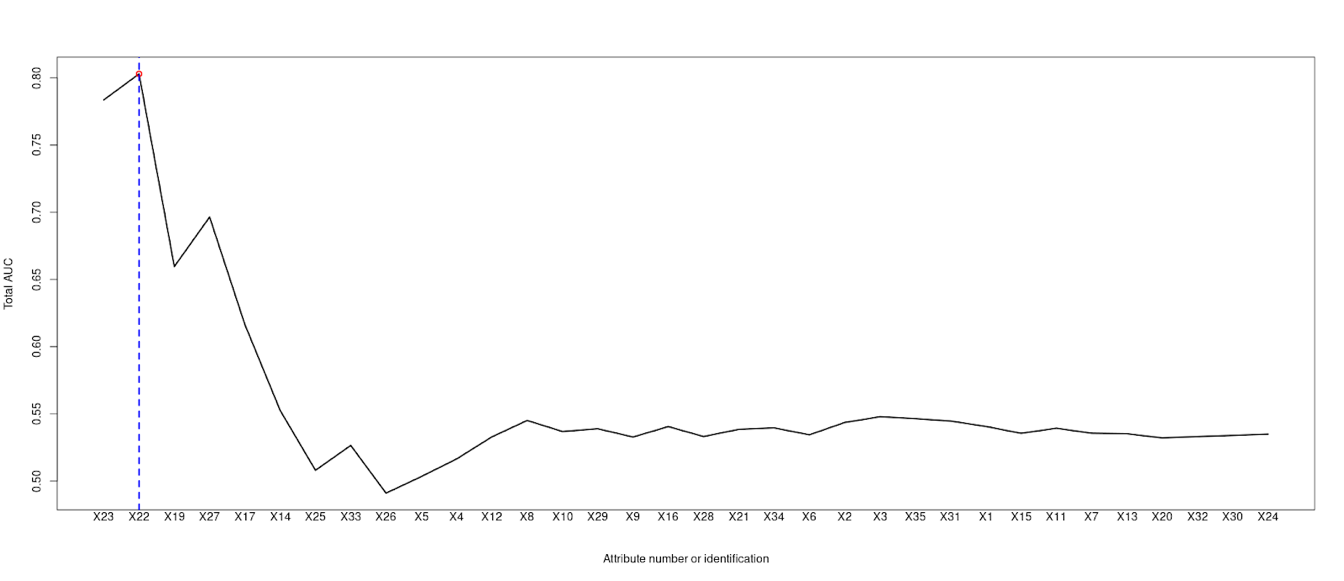

Legend: AUC: Area Under the Curve, X1: Anguish perception, X2: Anguish unpleasantness, X3: Bladder fullness perception, X4: Bladder fullness unpleasantness, X5: Cold perception, X6: Cold unpleasantness, X7: Dyspnea perception, X8: Dyspnea unpleasantness, X9: Muscle fatigue perception, X10: Muscle fatigue unpleasantness, X11: Gastric fullness perception, X12: Gastric fullness unpleasantness, X13: Heat perception, X14: Heat unpleasantness, X15: Heartbeat perception, X16: Heartbeat unpleasantness, X17: Hunger perception, X18: Hunger unpleasantness, X19: Itching perception, X20: Itching unpleasantness, X21: Muscle soreness perception, X22: Muscle soreness unpleasantness, X23: Nausea perception, X24: Nausea unpleasantness, X25: Deep pressure pain perception, X26: Deep pressure pain unpleasantness, X27: Respiratory frequency perception, X28: Respiratory frequency unpleasantness, X29: Sleep perception, X30: Sleep unpleasantness, X31: Thirst perception, X32: Thirst unpleasantness, X33: Visceral pain perception, X34: Visceral pain unpleasantness.

**Supplementary Figure S3.** Receiver-operating characteristic curve for stepwise area under the curve reduction.

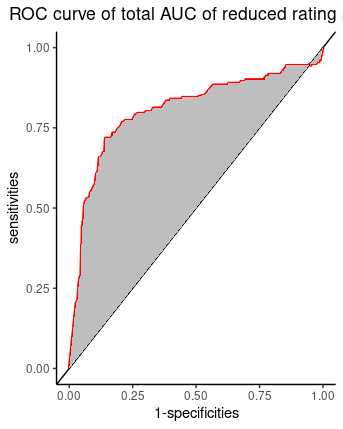

Legend: ROC: Receiver-operating characteristic, AUC: Area Under the Curve.

**Supplementary Figure S4.** Item response theory (IRT) information curves before and after item reduction.

S1a. Test information and standard	S1b. Test information and standard

errors before item reduction	erros after item reduction

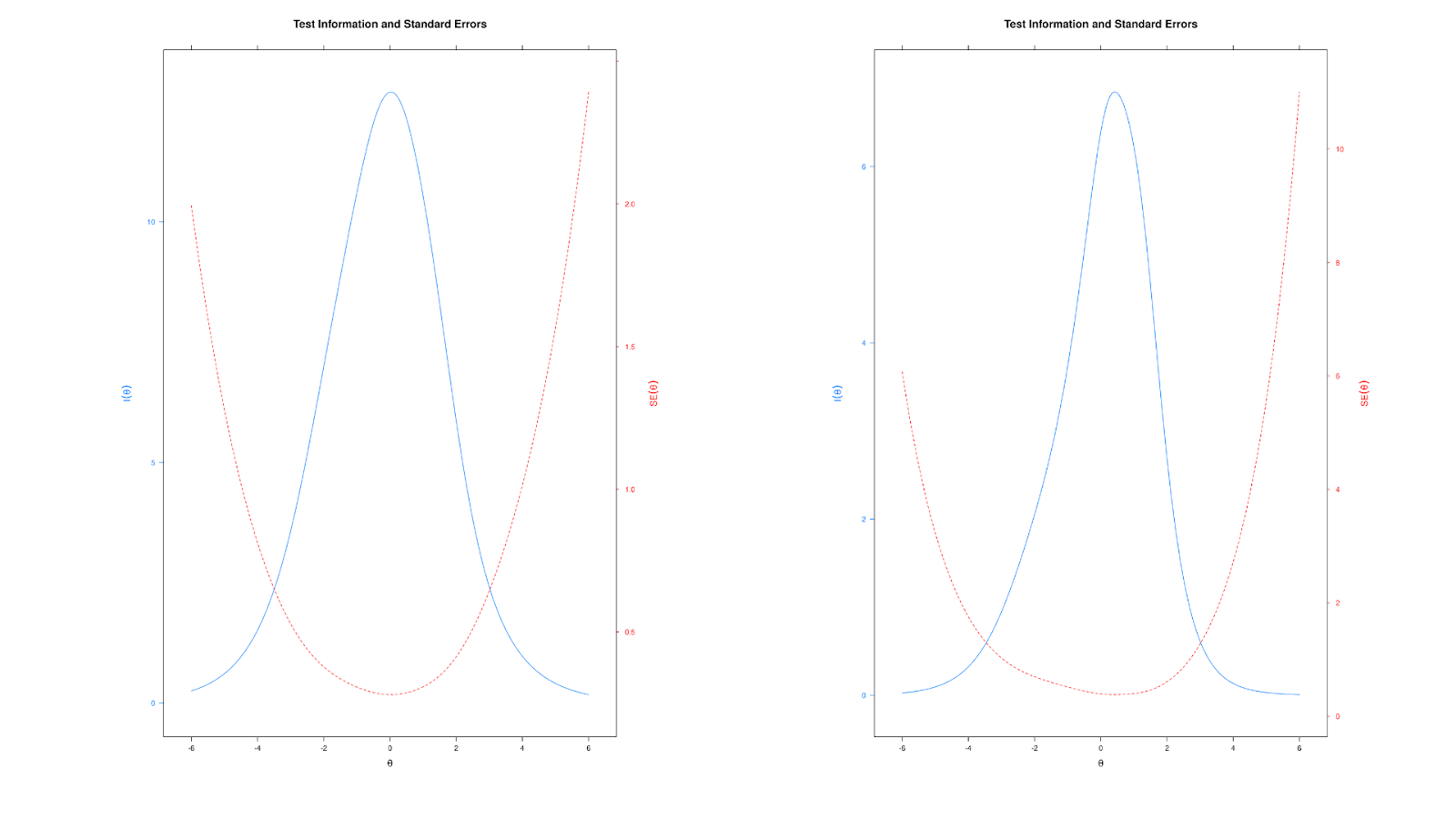

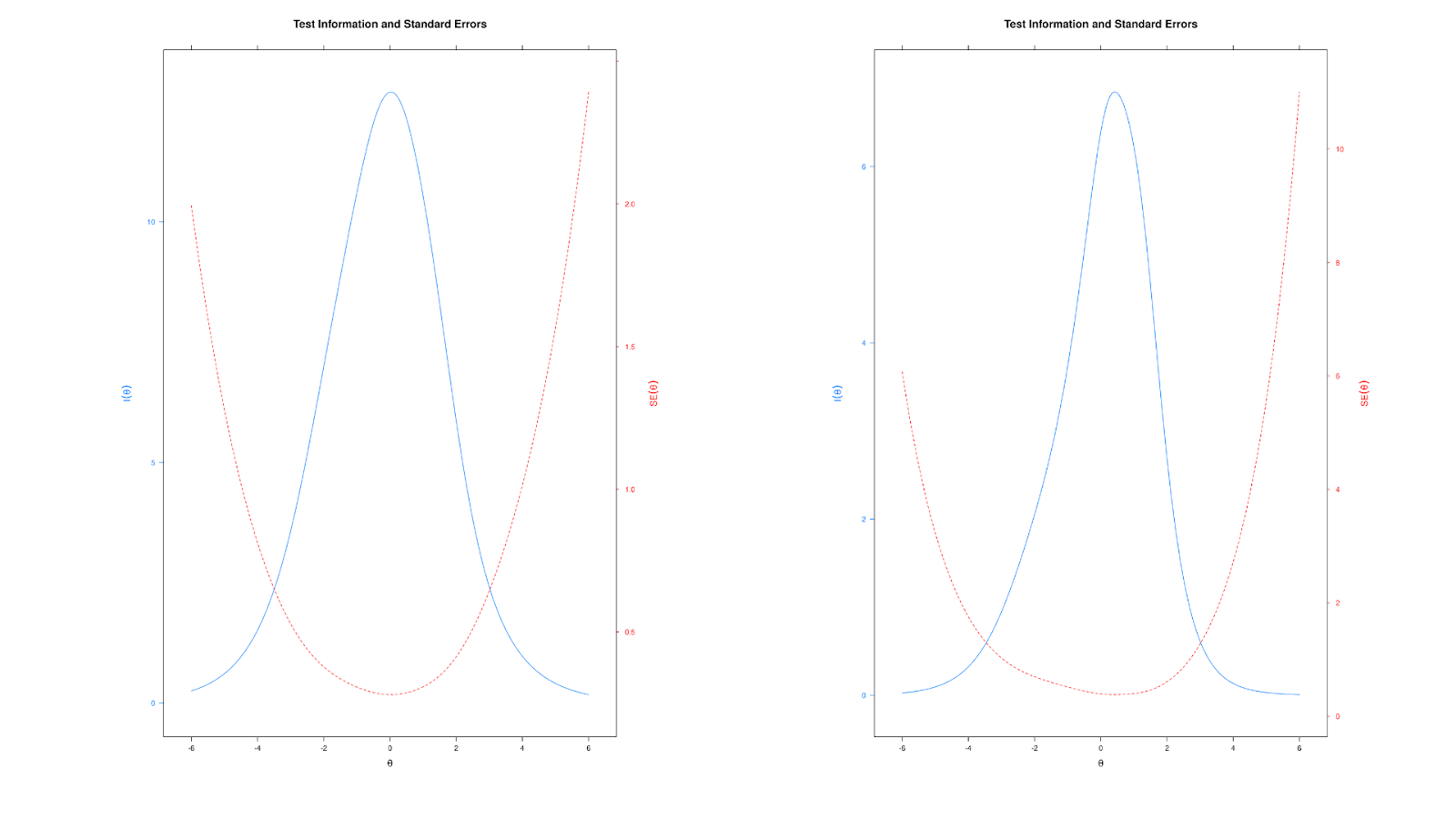

**Supplementary Table S4.** Item response theory (IRT)’s goodness-of-fit before and after item reduction.

| **Fit statistics** | **Before item reduction** | **After item reduction** |
| --- | --- | --- |
| M2 | 3,360 | 102 |
| df | 493 | 25 |
| *P* | *P* < 0.001 | *P* < 0.001 |
| RMSEA | 0.104 (0.101, 0.107) | 0.075 (0.06, 0.09) |
| SRMSR | 0.091 | 0.071 |
| TLI | 0.760 | 0.919 |
| CFI | 0.775 | 0.942 |

Legend: RMSEA: root mean square error of approximation, SRMSR: standardized root mean square residual, TLI: Tucker-Lewis Index, CFI: comparative fit index.

**Supplementary Figure S5.** Embodied Interoception Questionnaire (Intero-10).

**Introduction:** We can all perceive different sensations in our everyday lives, like hot, cold, pain, hunger or thirst. These sensations are personal and vary from one person to another. We want to know to what extent you perceive these sensations from inside of your body and / or to what extent some of them may bother you when they are present. For each question, you will see a line that indicates the increase of intensity from the left (lower intensity) to the right (higher intensity). The measuring scale ranges from "I do not perceive it at all / I am not bothered at all" on the leftmost position, to "I perceive it at the maximum possible intensity / It bothers at the maximum possible intensity" in the rightmost position. Please mark on the line what the intensity is for each question.

1) To what extent do you perceive your heart/chest palpitations in your daily life?

I do not perceive it at all____________________________________________________ I perceive it at the maximum possible intensity

2) To what extent do you perceive the feeling of being hot in your daily life?

I do not perceive it at all____________________________________________________ I perceive it at the maximum possible intensity

3) To what extent do you perceive the feeling of itch in your daily life?

I do not perceive it at all____________________________________________________ I perceive it at the maximum possible intensity

4) To what extent do you perceive the feeling of breathlessness in your daily life?

I do not perceive it at all____________________________________________________ I perceive it at the maximum possible intensity

5) To what extent do you to perceive whether you have slept well or not in your daily life?

I do not perceive it at all____________________________________________________ I perceive it at the maximum possible intensity

6) To what extent do you perceive the feeling of muscle fatigue in your daily life?

I do not perceive it at all____________________________________________________ I perceive it at the maximum possible intensity

7) To what extent do you perceive the feeling of anguish in your daily life?

I do not perceive it at all____________________________________________________ I perceive it at the maximum possible intensity

8) To what extent does it bother you to feel heart/chest palpitations in your daily life?

I am not bothered at all____________________________________________________ It bothers at the maximum possible intensity

9) To what extent does it bother you to feel breathless in your daily life?

I am not bothered at all____________________________________________________ It bothers at the maximum possible intensity

10) To what extent does it bother you to feel nauseous in your daily life?

I am not bothered at all____________________________________________________ It bothers at the maximum possible intensity

**Supplementary Table S5.** Score development classification after item reduction.

| **Category** | **Percentiles** | **Patient count** | **Score values** |
| --- | --- | --- | --- |
| Low | 0 to 3.33 | 125 | 0 to 4.25 |
| Medium | 3.34 to 6.67 | 124 | 4.26 to 5.76 |
| High | 6.68 to 10 | 124 | 5.77 to 10 |

**Supplementary Table S6:** Test-retest reliability in short- and long-term.

|  | **Short term (3h)**  **n (56)** |  | **Long term (30d)**  **n (44)** |  |
| --- | --- | --- | --- | --- |
|  | **ICC** | **CI95%** | **ICC** | **CI95%** |
| Heartbeat perception | 0.84 | (0.73-0.90) | 0.60 | (0.28-0.78) |
| Heat perception | 0.88 | (0.80-0.93) | 0.61 | (0.30-0.78) |
| Itch perception | 0.83 | (0.71-0.90) | 0.73 | (0.50-0.85) |
| Dyspnea perception | 0.86 | (0.74-0.92) | 0.55 | (0.18-0.75) |
| Sleep perception | 0.93 | (0.88-0.95) | 0.73 | (0.51-0.85) |
| Muscle fatigue perception | 0.92 | (0.87-0.95) | 0.80 | (0.63-0.89) |
| Anguish perception | 0.89 | (0.81-0.93) | 0.63 | (0.33-0.80) |
| Heartbeat unpleasantness | 0.85 | (0.73-0.91) | 0.70 | (0.43-0.83) |
| Dyspnea unpleasantness | 0.86 | (0.76-0.91) | 0.81 | (0.66-0.90) |
| Nausea unpleasantness | 0.90 | (0.83-0.94) | 0.83 | (0.68-0.90) |

Legend: ICC: intraclass correlation coefficient, h: hours, d: days.

**Supplementary Table S7.** Content validity coefficient for the introduction and 10 questions from Intero-10 accessed by potential users.

|  | **Clarity** | | | **Relevance** | | |
| --- | --- | --- | --- | --- | --- | --- |
|  | **Mean** | **CVC *i*** | **CVC *t*** | **Mean** | **CVC *i*** | **CVC *t*** |
| Introduction | 3.8 | 0.95 | 0.94 | 4.0 | 1.00 | 0.99 |
| Heartbeat perception | 4.0 | 1.00 | 0.99 | 3.8 | 0.80 | 0.94 |
| Heat perception | 4.0 | 1.00 | 0.99 | 4.0 | 0.95 | 0.99 |
| Itch perception | 4.0 | 1.00 | 0.99 | 4.0 | 1.00 | 0.99 |
| Dyspnea perception | 4.0 | 1.00 | 0.99 | 4.0 | 1.00 | 0.99 |
| Sleep perception | 3.8 | 0.95 | 0.94 | 4.0 | 0.95 | 0.99 |
| Muscle fatigue perception | 4.0 | 1.00 | 0.99 | 4.0 | 0.95 | 0.99 |
| Anguish perception | 4.0 | 1.00 | 0.99 | 4.0 | 0.95 | 0.99 |
| Heartbeat unpleasantness | 4.0 | 1.00 | 0.99 | 4.0 | 1.00 | 0.99 |
| Dyspnea unpleasantness | 4.0 | 1.00 | 0.99 | 4.0 | 0.95 | 0.99 |
| Nausea unpleasantness | 4.0 | 1.00 | 0.99 | 4.0 | 0.95 | 0.99 |

Legend: CVC *i* : initial content validity coefficient, CVC *t*: total content validity coefficient.
